# A Meta-Analytic Investigation of Grey Matter Differences in Anorexia Nervosa and Autism Spectrum Disorder

**DOI:** 10.1101/2022.03.17.22272346

**Authors:** Michelle Sader, Justin H. G. Williams, Gordon D. Waiter

## Abstract

Recent research reports Anorexia Nervosa (AN) to be highly dependent upon neurobiological function. Some behaviours, particularly concerning food selectivity are found in populations with both Autism Spectrum Disorder (ASD) and AN, and there is a proportionally elevated number of anorexic patients exhibiting symptoms of ASD. We performed a systematic review of structural MRI literature with the aim of identifying common structural neural correlates common to both AN and ASD. Across 46 ASD publications, a meta-analysis of volumetric differences between ASD and healthy controls revealed no consistently affected brain regions. Meta-analysis of 23 AN publications revealed increased volume within the orbitofrontal cortex and medial temporal lobe, and adult-only AN literature revealed differences within the genu of the anterior cingulate cortex. The changes are consistent with alterations in flexible reward-related learning and episodic memory reported in neuropsychological studies. There was no structural overlap between ASD and AN. Findings suggest no consistent neuroanatomical abnormality associated with ASD, and evidence is lacking to suggest that reported behavioural similarities between those with AN and ASD are due to neuroanatomical structural similarities.

## 1.0 INTRODUCTION

### 1.1 AN OVERVIEW OF AN AND ASD

Anorexia nervosa (AN) is a severe and multifactorial eating disorder characterised by a pathological fear of putting on weight and a distorted body-image. This psychiatric condition results in extremely low body weight in the context of individual age, sex, and overall development. AN affects 8 in 100,000 people, and has the highest mortality rate of any psychiatric illness at 20%^1,2^. In tandem, ASD is a neurodevelopmental condition predominantly characterised by a wide range of pathological social and behavioural impairments. Social deficits reflect difficulty expressing reciprocal verbal and non-verbal behaviour, but patients may also show rigidity in behaviour and hypersensitivity to external stimuli^3^. Significant financial^4^ and emotional^5^ burdens, as well as increased reports of the condition have sparked global concern^6^.

A growing body of work reports commonalities in the cognitive profiles and reported behavioural symptomatology between those with AN and ASD^7^. Baron-Cohen (2013) discusses the existence of multiple factors connecting anorexic and autistic traits, such as impairment in empathy as well as food selectivity and difficulty eating in social settings^8^. A significant commonality expressed in both conditions relates to behavioural and attitude-based rigidity with a particular focus on food or weight in AN^9^. A meta-analysis by Westwood et al., (2016)^10^ assessed cognitive flexibility in those with AN and ASD via the Wisconsin Card Sorting Test (WCST)^10,11^. While there were no reports concerning effect of diagnosis on performance, ASD and AN groups made significantly higher perseverative errors. Moreover, both clinical groups share behavioural parallels related to difficulty in empathizing or relating to others. Individuals with AN may show impaired empathy by employing most of their attentional resources internally and engaging in social comparison behaviour^12,13^. Impairment in executive function, central coherence and empathy has also been widely noted in those with autism^14-19^. A genome-wide association (GWAS) study involving 46,861 participants evaluated the relationship between Emotional Quotient (EQ) performance and risk for neuropsychological conditions such as AN, ASD and schizophrenia. Significant negative correlations were found between EQ performance and ASD, as well as a positive correlation between EQ performance and genetic risk for anorexia nervosa^17^.

### 1.2 NEUROIMAGING OVERLAP

Beyond cognitive measurements, structural and functional imaging suggest that AN and ASD could share condition-related brain regions. A variety of previous research has reported aberrant functionality in both AN and ASD within social and appetitive centres such as the amygdala, cerebellum, insula, cingulate cortex, fusiform gyrus and superior temporal gyrus^20^^-^^24^. The anterior cingulate cortex (ACC), insula and cerebellum, which are associated with emotional learning, have been reported to be discrepant in both anatomical size and structural connectivity when compared to controls^25-29^. The insula, hippocampus and cerebellum are three key regions of the brain reported to be structurally altered in both conditions^27^,^30^^-^^34^.

Abnormal neuroanatomical structure and function reported in AN and ASD is consistent with a growing body of work investigating emotional and cognitive disorder aspects, such as food selectivity, behavioural and attitude-based rigidity as well as impaired Theory of Mind^10,16,35^. For example, increased activity of ACC has been noted to correlate with unpleasantness and pain processing, and has been associated with psychosocial behaviours seen in AN, such as impaired emotional awareness and reasoning as well as increased alexithymia scores^36,37^. Increased volume of the orbitofrontal cortex (OFC) has been associated with food avoidance in AN^38^. Other studies report reduced attention to facial stimuli and decreased understanding of other’s emotions in AN^39,40^.

Voxel-based morphometry (VBM) is a widely utilised neuroimaging technique that investigates focal differences in neuroanatomy through segmentation, normalisation and smoothing of tissues into grey matter, white matter, and cerebrospinal fluid^41^. VBM is particularly amenable to meta-analytic reviews as the method corrects for individual differences in structural anatomy^42^, and may be utilised across groups of individuals.

Two previous meta-analyses have evaluated neuroanatomical structure in AN using VBM^28,29^. These studies both show findings of decreased volume within the cingulate cortex, hypothalamus and frontal/parietal lobes, but have not considered some more recent VBM findings^38,43,44^. In ASD, multiple VBM meta-analyses have examined condition-related neurocorrelates, reporting widespread structural alteration in white and grey matter volume, including the frontal/parietal lobes, medial temporal lobe, lateral occipital lobe, bilateral cerebellum, middle temporal gyrus, angular gyrus, fusiform gyrus, insula, right ACC, posterior cingulate cortex, caudate, amygdala and precuneus^45-50^. However, individual studies examining the neuroanatomy associated with ASD often contain small sample sizes of less than 20 per cohort^51-61^. Additionally, no recent meta-analytic work has incorporated research from 2017 onwards. Despite previous meta-analytic VBM research in AN and ASD respectively, there are additionally no structural studies investigating overlap in brain structure, and existing review literature focuses on self-reported questionnaire measurements rather than MRI modalities^10,62^. One VBM study of note conducted by Björnsdotter *et al*. (2018)^63^ evaluated correlations between neuroanatomical structure and scores on the Autism Quotient (AQ) questionnaire in those with AN. Compared with controls, those with AN exhibited decreased volume in the bilateral superior temporal sulcus (STS), with the left STS negatively correlating with AQ scores^63^. However, this work consists of the limitation that use of the AQ questionnaire in a non-ASD population only serves as a behavioural marker, and cannot be considered a diagnostic factor of AN. In summary, AN/ASD-related neuroimaging research in the past decade has grown extensively and implicated possible regions of overlap, but requires further investigation.

We collated AN and ASD VBM literature utilising large-cohort MRI data to quantify, identify and elaborate upon regions of the brain associated with AN and ASD. While cross-sectional studies have limited capacity to understand the causal or longitudinal nature of neuroanatomical structure in AN or ASD, separately evaluating distinct age subgroups compared with the combined dataset will provide information regarding what specific regions of the brain are impacted within distinct age ranges, and whether an existing overlap in affected structure will change according to age.

## 2.0 METHODOLOGY

### 2.1 SELECTION OF LITERATURE

Literature was selected using SCOPUS, PubMed, and Web of Science (WOS). From the used key terms, PubMed revealed 604 publications regarding states of hunger and anorexia [*search criteria*: ((Anorexia OR Anorex* OR Anorexia nervosa OR AN OR Eating Disorder) AND (VBM OR voxel based morphometry AND structural magnetic resonance imaging OR sMRI))]. SCOPUS identified 3139 publications reflecting AN research. Lastly, WOS identified 2498 papers regarding AN or states of undereating (n_total_=6,241). For our ASD literature selection, PubMed revealed 1503 publications relating to ASD [*search criteria*: ((ASD OR Autis* OR Asperger* OR PDD OR Pervasive Developmental Disorder) AND (VBM AND voxel based morphometry AND structural magnetic resonance imaging OR sMRI))]. SCOPUS identified 2507 documents under the utilised search criteria and Web of Science identified 45 publications (n_total_=4,055) Initial inclusion criteria involved the presence of key phrases utilised in search. Exclusion criteria were as follows: 1.) Publications older than 20 years, with 2000 as the cut-off range for literature selection; 2.) Animal studies, 3.) Publications lacking case-controls; 4.) Meta-analyses; 5.) No VBM analysis, or results not reported in Talairach or MNI coordinates. Within the AN literature selection, individual studies fitting search criteria were additionally collected from identified meta-analyses (n=4). In sum, 46 ASD and 23 AN papers included coordinates and were selected for final analysis with 5,236 total subjects (nASD=2062 vs. nHC=2044; nAN=536 & nRAN=37 vs. nHC=557) (*Table 1*.). 780 coordinates were collected from selected publications. (*Figure 1; Supplementary Table 1*.).

**Table 1.**
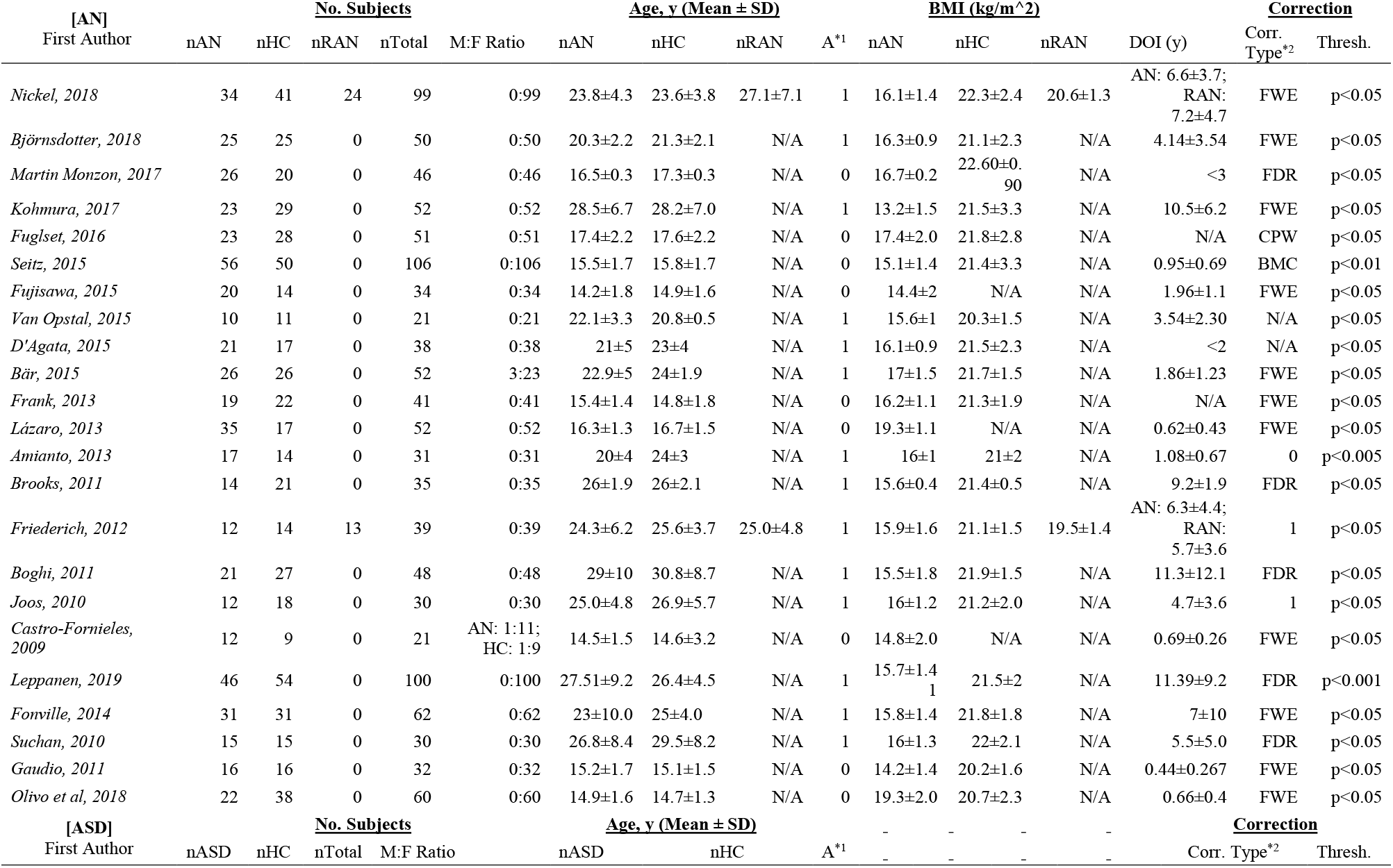
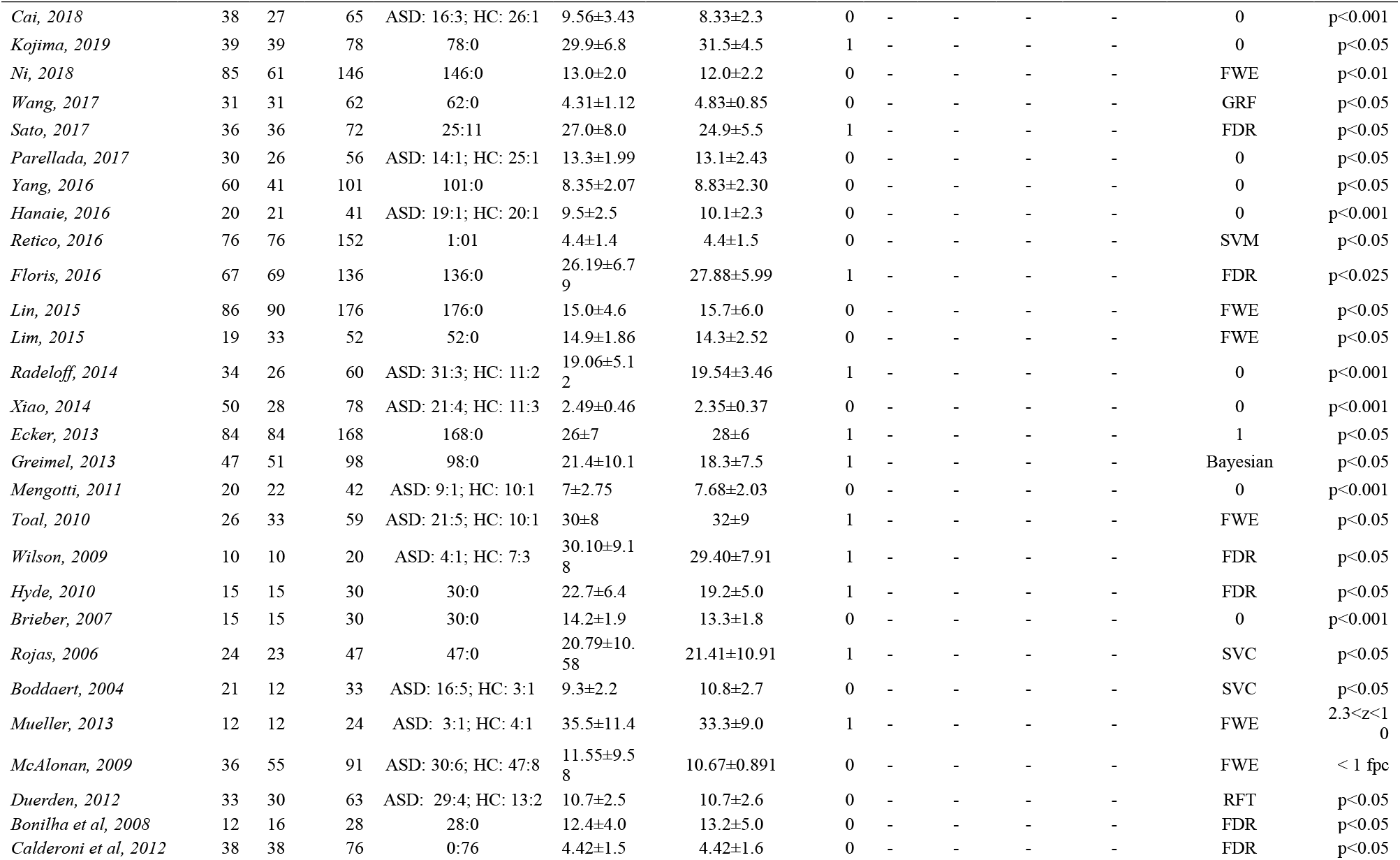
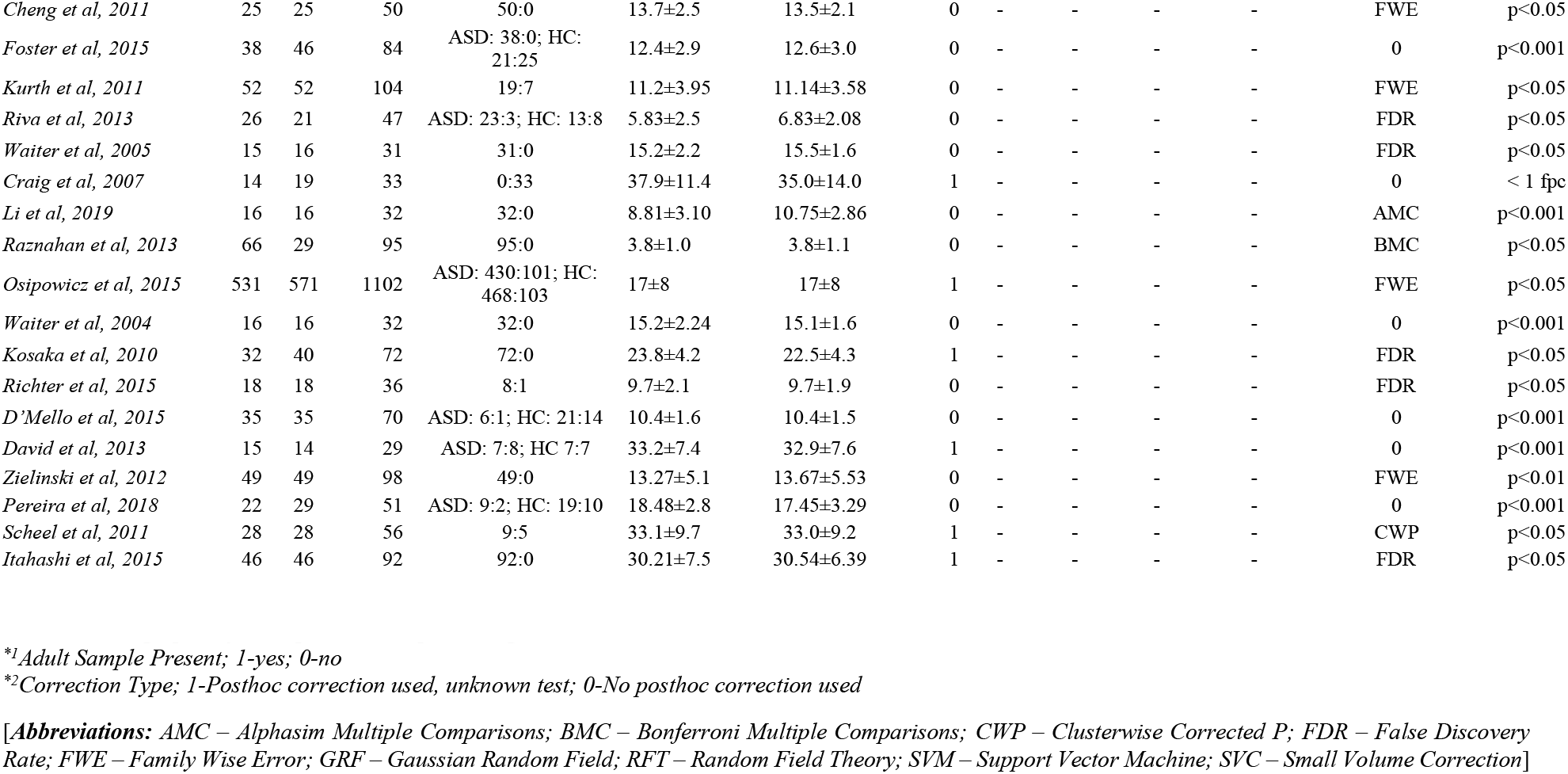
Demographics for the publications pertaining to aberrant brain volume in both AN and ASD data.

**Figure 1.**
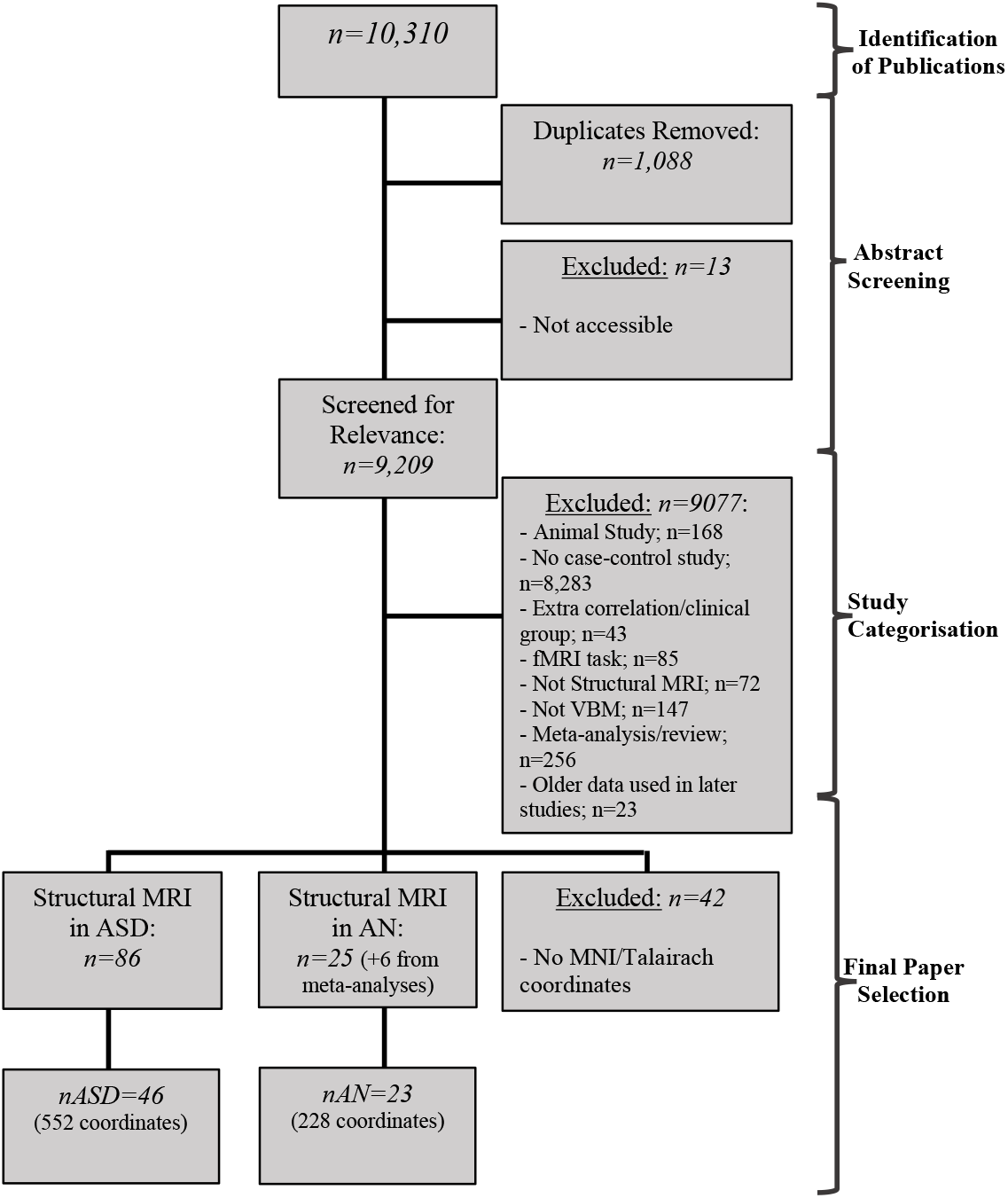
Flowchart depicting the identification, screening, and acquisition of data.

### 2.2 VOXEL-BASED MORPHOMETRY AND ALE ANALYSIS

Coordinates were obtained from the AN and ASD literature searches in Montreal Neurological Institute (MNI) and Talairach space. Coordinates were respectively incorporated into GingerALE, an Activation Likelihood Estimate (ALE) meta-analysis software^64^. GingerALE converted coordinates obtained in Talairach Spaces to MNI spaces using the *icbm2tal* transform. P thresholds for individual analyses were corrected for family-wise error (FWE) and set to p<0.05. As the GingerALE software calculates studies reporting increase and decrease clusters separately, positive and negative volume differences in each clinical group were separately analysed and subsequently pooled together. Once whole-brain data for clinical groups independent of age was combined, further analyses were conducted to analyse neuroanatomical structure in the adult (21+ y old) literature and identify if findings are significantly different than those from child (0-20 y old) studies. Data was then pooled to observe respective increases and decreases dependent on age. There was insufficient literature to compare child-only studies in AN with our combined dataset.

Once age-dependent clusters were identified by the GingerALE software, results were compared between AN and ASD for a final conjunction analysis to investigating potential overlap in volume abnormalities within age-dependent subgroups. FWE-corrected clusters produced by GingerALE were visualised on *Mango*^65^. Using an MNI template file, the output FWE-corrected clusters generated from GingerALE are inserted as an overlay to visualise regions to identify any overlap in affected regions. In order to prevent conflation of AN, ASD and pooled sample data, any conjunction analyses on reported increase/decrease results were conducted separately. Executing separate analyses for increase and decrease data allows a given result value to retain its significance amongst scientific literature, as well as prevents duplication of sample sizes. Subject data was uncorrected under a threshold of p<0.05.

## 3.0 RESULTS

In summary, 70 papers^21,38,43,44,48,51-61,63,66-115^ included coordinates and were selected for final analysis with 5,484 total subjects (nAN=562 & nRecovered AN (RAN)=37 vs. nHC=584; nASD=2164 vs. nHC=2137) (*Table 1*.). 786 coordinates were collected from the selected publications. (*Figure 1*.). This study also included work related to the ABIDE II dataset, which consists of large-scale health and imaging cohort data of those with ASD relative to controls. To prevent conflation of ABIDE II data, the publication with the largest ABIDE II sample size to date was included for analysis^107^. Collected literature in AN was female-predominant, with only two publications recruiting male subjects^69,77^. Meanwhile, literature in ASD was strongly-male predominant, with two publications recruiting female-only subjects^58,101^. Mean participant age was 21.0±4.22 for AN subjects (RAN=26.05±6) and 21.5±3.18 for HCs, and the mean age in ASD publications was 15.6±9.4 for ASD and 15.6±9.1 for HCs.

### 3.1 VOLUMETRIC INCREASE AND DECREASE IN ASD

Despite a multitude of previous brain regions being reported in VBM literature (*Figure 2*.), no significant clusters indicating positive or negative volume differences were found when testing all ASD data at p<0.001 or under FWE correction at p<0.05, both dependent and independent of age.

**Figure 2.**
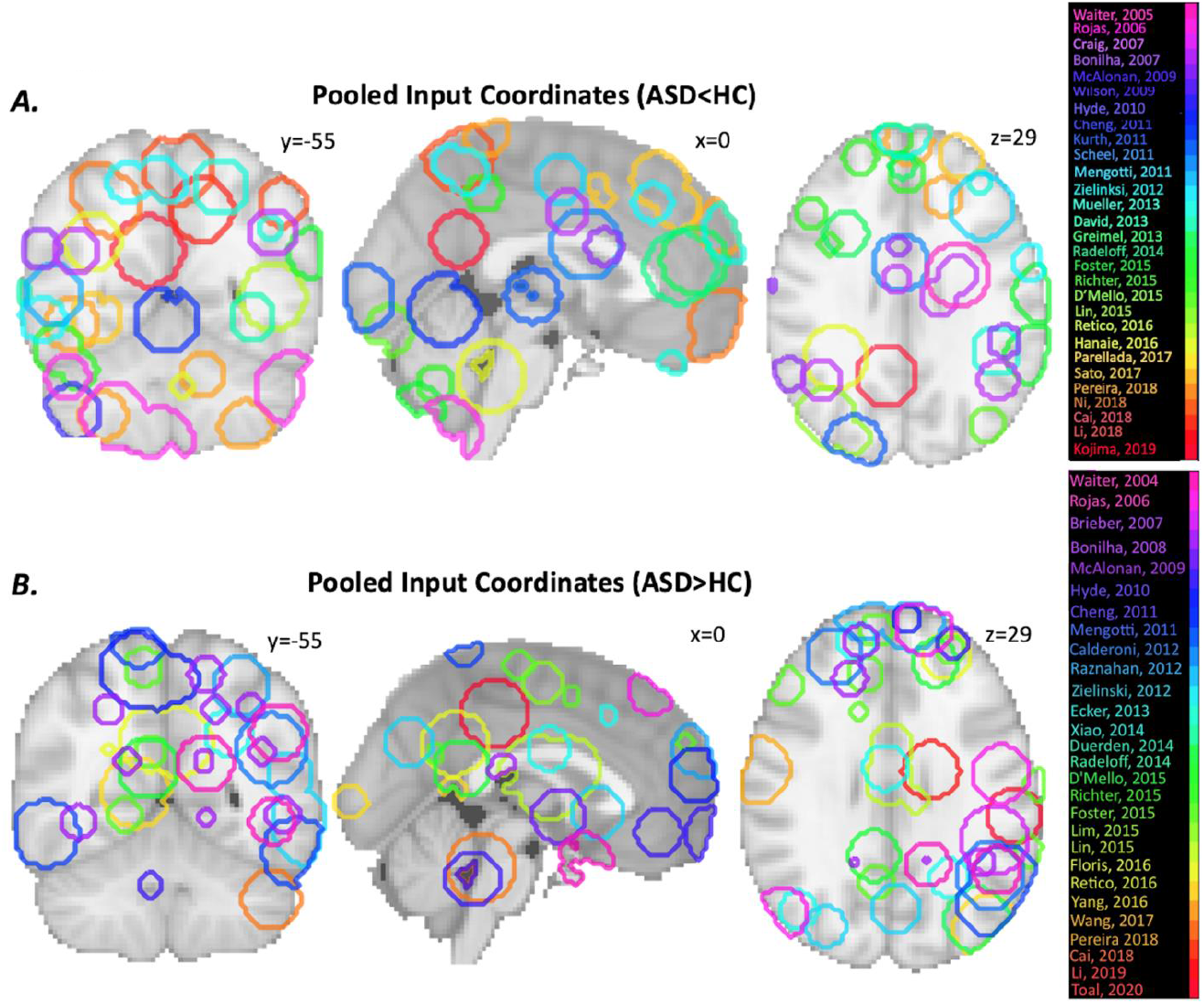
Input coordinates delineated via Nilearn (Python) demonstrate the extensive diversity of implicated structural decrease (**A**) and increase (**B**) across case-control ASD literature. As not all publications are visible from one brain slice, MNI coordinates represent slices of the brain with the majority of visible clusters from available literature.

### 3.1 VOLUMETRIC INCREASE AND DECREASE IN AN

Combining all coordinates from AN papers published within the past 20 years revealed significant differences in grey matter volume within a number of regions. Results of the GingerALE analysis identified three clusters of increased volume primarily located within the left and right medial temporal lobes. The largest cluster (28.72cm^3^) with a centre at x=29, y=-21, z=-22 was largely located within the right parahippocampal gyrus. A second cluster (16.12cm^3^) was located within the left uncus and amygdala with a centre at x=-32, y=-7, z=-41.A third cluster was (8.58cm^3^) was located on the left OFC and surrounding medial frontal gyrus (MFG) with a centre at x=-7, y=38, z=-25 (*Table 2*.). While areas of increased volume were predominantly surface-based, they also included subcortical structures such as the amygdala and hippocampus (*Figure 3*.*1*.). Results additionally identified a singular cluster reflecting volume reduction in AN subjects compared with a typical population primarily located within the right cingulate gyrus (7.29cm^3^) with a centre at x=10, y=-28, z=44, (*Table 2*.). The precuneus, lateral prefrontal cortex and paracentral lobule additionally demonstrated reductions in volume (*Figure 3*.*2*.).

**Table 2.** Decreased GMV in AN participants relative to healthy controls (AN < HC) including all AN studies (n=23). Data is presented in order of significance, with coordinate sets in bold comprising of cluster centres.

| [ALL<br>DATA]<br>AN>HC | Region | BA | MNI<br>Coordinates |  |  | Volume<br>(mm<sup>3</sup>) | ALE<br>score | P | Z |
| --- | --- | --- | --- | --- | --- | --- | --- | --- | --- |
|  |  |  | x | y | z |  |  |  |  |
| **C1** | **R Parahippocampal Gyrus** | **35** | **28.8** | **-21.2** | **-21.5** | **28,720** | **0.0078** | **9.12E-04** | **3.12** |
| **C1** | **R Parahippocampal Gyrus** | **28** | **29** | **-24.3** | **-16** |  | **0.0051** | **3.50E-03** | **2.7** |
| **C2** | **L Uncus** | **36** | **-32** | **-7** | **-41** | **16,123** | **0.009** | **2.22E-04** | **3.51** |
| **C2** | **L Parahippocampal Gyrus** | **N/A** | **-27** | **-16** | **-19** |  | **5.03E-04** | **2.94E-02** | **1.89** |
| **C3** | **L Orbitofrontal cortex (MFG)** | **11** | **-6.7** | **37.7** | **-25.3** | **8,584** | **9.54E-04** | **2.10E-02** | **2.03** |
| AN<HC |  |  |  |  |  |  |  |  |  |
| **C1** | **R Cingulate Gyrus** | **31** | **10** | **-28** | **44** | **13,152** | **0.024** | **5.25E-07** | **4.89** |
| **C1** | **L Cingulate Gyrus** | **31** | **-4** | **-40** | **32** |  | **0.016** | **7.98E-05** | **3.78** |
| **C1** | **L Precuneus** | **7** | **-4** | **-56** | **52** |  | **0.015** | **1.39E-04** | **3.63** |
| **C1** | **L Paracentral Lobule** | **31** | **-2** | **-16** | **46** |  | **1.50E-02** | **1.46E-04** | **3.62** |
| **C1** | **R Precuneus** | **7** | **6** | **-58** | **44** |  | **1.30E-02** | **5.38E-04** | **3.27** |
| **C1** | **R Cingulate Gyrus** | **31** | **8** | **-44** | **40** |  | **0.012** | **8.83E-04** | **3.13** |
| **C1** | **R LPFC (Middle Frontal Gyrus)** | **6** | **24** | **-6** | **52** |  | **0.012** | **8.97E-04** | **3.12** |
| **C1** | **R LPFC (Middle Frontal Gyrus)** | **6** | **26** | **-2** | **50** |  | **1.90E-02** | **1.01E-03** | **3.09** |
| **C1** | **R LPFC (Middle Frontal Gyrus)** | **6** | **22** | **-6** | **58** |  | **1.10E-02** | **1.64E-03** | **2.94** |
| **C1** | **R Sub-Gyral** | **6** | **30** | **8** | **54** |  | **0.011** | **2.02E-03** | **2.87** |
| **C1** | **L Cingulate Gyrus** | **31** | **-4** | **-26** | **44** |  | **0.01** | **2.97E-03** | **2.75** |
| **C1** | **L Cingulate Gyrus** | **24** | **-10** | **-16** | **38** |  | **1.00E-02** | **3.17E-03** | **2.73** |
| **C1** | **R LPFC (Middle Frontal Gyrus)** | **6** | **26** | **8** | **64** |  | **1.00E-02** | **3.91E-03** | **2.65** |
| **C1** | **R Cingulate Gyrus** | **31** | **22** | **-18** | **48** |  | **9.00E-03** | **4.55E-03** | **2.6** |

[Abbreviations: L – left; R – right; LPFC – Lateral prefrontal cortex; MFG – Medial frontal gyrus; BA – Brodmann Area; C - Cluster]

**Figure 3.**
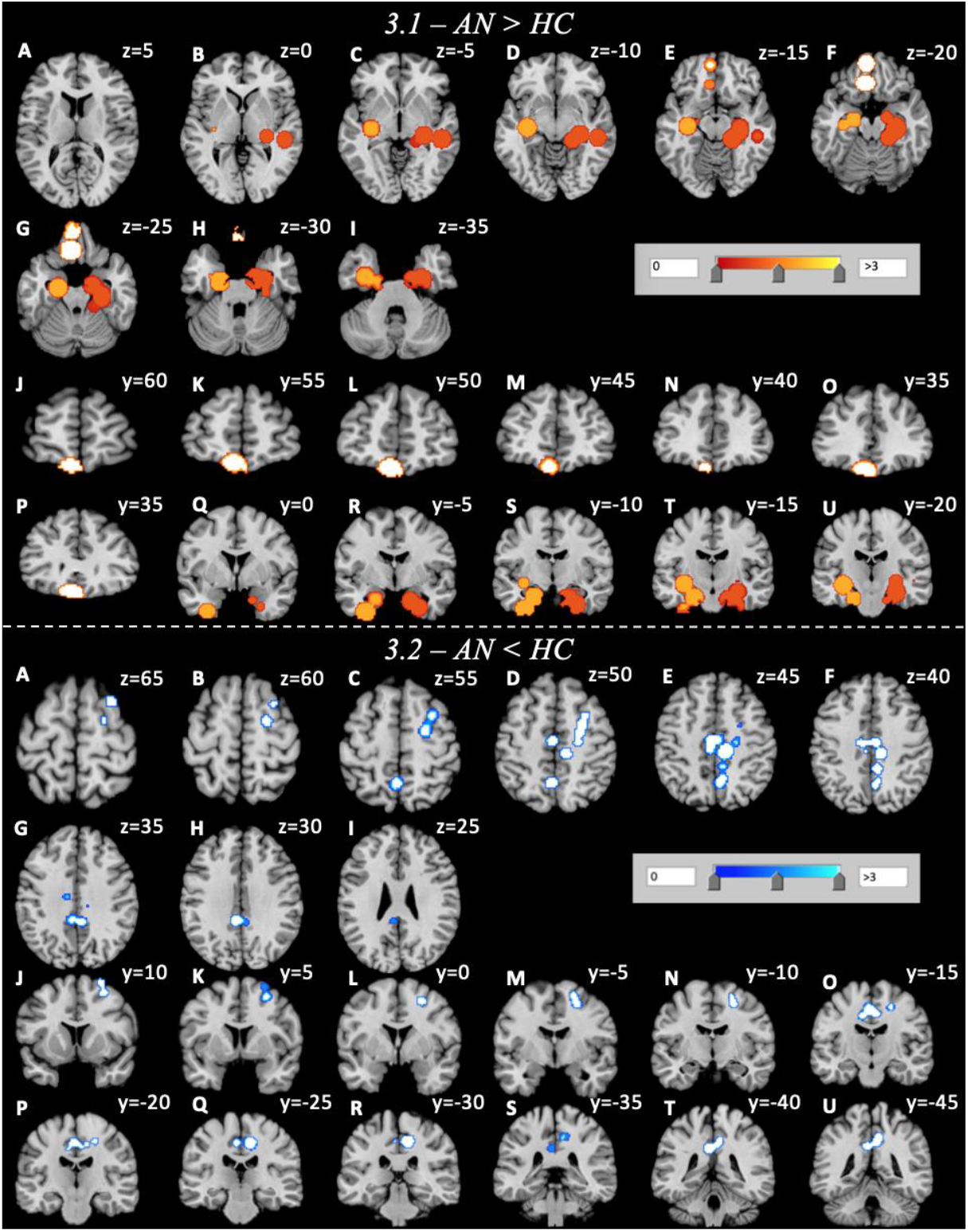
Clusters of volume increase (**3.1)** and decrease (**3.2**) in AN participants relative to healthy controls. Volumetric increase in those with AN spans from z=5 to z=-35, y=50 to y=20/y=0 to y=-20 and x=-30 to x=30 in axial (**A-I**) and coronal (**J-U**) orientations. Volumetric decrease in those with AN spans from z=65 to z=20, y=10 to y=-45 and x=-5 to x=25 in axial (**A-I**) and coronal (**J-U**) orientations.

Repeating analysis of AN data after removal of child publications altered the findings in some respects, and identified significant additional differences when compared with all collated data (*Supplementary Figure 1*). As with the all-data analysis, analysis revealed three large clusters indicating increase in volume. The largest cluster (14cm^3^) with a centre at x=12, y=-38, z=52 was predominantly located within the right paracentral lobule. The second cluster (12.18cm^3^) was located within the left anterior cingulate cortex (L ACC). The third cluster was found within the right lateral prefrontal cortex and was centered at x=38, y=52, z=20. Similar to results when all data was included, removal of child data additionally revealed a large cluster of decreased volume within the right cingulate gyrus, with slightly increased volume (13.15cm^3^ to 17.77cm^3^) (*Supplementary Table 2*). In contrast to analysing all AN data, the adult-only clusters indicated that peaks of significance were more focused within the ACC as opposed to the OFC observed in the entire dataset.

## 4.0 DISCUSSION

This systematic review and meta-analysis presents the first analysis of VBM literature comparing AN and ASD with 70 publications contributing to the study. Inspecting the diverse range of implicated regions from publications identified in this review (*Figure 2*.), we found no significant clusters indicating regions with common or consistent volumetric increase or decrease when comparing ASD relative to HCs. Separating data by age also showed no significant common volumetric differences in those with ASD relative to controls across the literature. In contrast, review of AN literature demonstrated marked effects in regions of the brain associated with executive function, memory consolidation, conflict monitoring and decision-making. Age-dependent analyses delineate further structural aberrance of the ACC when comparing adult and child cohorts.

Regarding our review of ASD VBM literature, a lack of condition-associated regions directly contradicts previously reported meta-analytic findings in ASD^45-50^. Previous reviews report disturbance within the cerebellum^45^, precuneus^50^, cingulate cortex^45^, fusiform gyrus^45^, fronto-temporal lobes^45^,^47-50^, parietal lobes^45,49^, occipital lobes^46^,^48-50^ and the basal ganglia^46-48^. However, it is pertinent to note the extreme diversity in associated brain regions and direction of effect. For instance, findings regarding the temporal lobe of the brain, implicated in the majority of previous VBM reviews, were discrepant in location and affected either the medial temporal lobe/gyrus (MTL/G)^45,48^, inferior temporal gyrus^50^ or temporal poles^49^. Additionally, such findings are unable to unilaterally confirm whether the temporal lobe is structurally increased^45,47,50^ or decreased^45,47,49^ in those with ASD relative to controls. Regional discrepancies pertaining to location and effect can also be identified within the cerebellum^45^ and frontal brain regions (i.e., middle frontal gyrus, prefrontal cortex)^48-50^. Further, some reviews note that volumetric abnormalities, particularly structural reduction, are dependent on age^47-49^, while other reviews either did not identify associations between age and regional volume^46^, or were assessing age-specific cohorts^46,50^.

A lack of consistent condition-associated brain regions in this meta-analysis may also be explained by high level of heterogeneity and comorbidity that occurs in ASD. For example, estimates of the frequency of intellectual impairment are highly vaired in ASD, ranging from 10 to 90% depending on sampling and diagnostic definitions^116^. Anxiety and ADHD are other commonly presented neurodevelopmental problems^117^. Thus, neurodevelopmental impairment may commonly occur in ASD which is not directly related to the behavioural syndrome that characterises ASD. Structural neuroimaging studies are cross-sectional and only identify associations rather than causative links between differences and condition. It is therefore possible that differences reported in previous studies reflect associations with non-specific neural correlates, rather than specific neural correlates which characterise and may play a causal role in the etiology of ASD. Altogether, the diverse array of implicated regions and contrasting findings regarding structural effect in previous ASD literature in tandem with characteristic heterogeneity of causes may explain the absence of consistent findings seen in this meta-analysis.

In contrast to the ASD analysis, a review of AN VBM literature provides an updated report demonstrating significantly altered neuroanatomy associated with the AN cohort compared to controls, and is the first quantitative analysis to report volumetric *increase* in AN^28,29^. Results from analysing 23 AN publications revealed common regions of increased volume in the parahippocampal gyrus, OFC and MTL/medial frontal gyrus (MFG). The OFC is tightly interconnected with the ACC which work together to modulate reward- and appetite-based learning^118,119^. This brain region plays important roles in regulating food intake and contains the site where the reward/value of taste is processed^118^. The OFC also integrates aversive forms of taste, and is subject to reinforcement association to positive and negative stimuli^119^. OFC-related predictions based on individual reward value may play an important role in those with AN, as food may become associated with a prediction of weight gain and negative body image, both of which become strongly aversive stimuli in AN. Our findings regarding increased or unusual OFC volume in AN have not been identified by previous VBM meta-analyses^28,29^. However, a growing number of authors identify increased OFC volume and cortical thickness, and suggest that the OFC may contribute to traits of food avoidance in AN^44,120,121^.

Abnormal parahippocampal gyrus volume has been previously reported in AN^122-124^ and serves important roles in episodic memory, including visual scene recognition, encoding, retrieval^125,126^, autobiographical memory formation^122,123,127,128^ and integration of social contexts^129,130^. Findings reflect a model presented by Riva *et al*. (2016)^131^ explaining how increased autobiographical memory for negative experiences in AN affects egocentric body memory and lead to distorted body image^131^. The parahippocampal cortex is important the computation of third-person or allocentric memories, both autobiographical and episodic^132^.

Decreased volume in AN relative to HCs was observed in the bilateral cingulate gyrus, right lateral prefrontal cortex (LPFC)/middle frontal gyrus, cuneus and paracentral cortex. In AN, reduction of the LPFC may be involved in the cognitive and motor aspects of inhibitory control as a means to maintain dietary restriction.

Analysis upon removal of child studies revealed distinct regions of abnormal neuroanatomy when compared with the entire AN cohort. While clusters of reduced volume were relatively unaffected, states of increased volume greatly differed between adult-only AN data and the collected dataset. The right superior/MFG and ACC were the most affected, with the ACC cluster not appearing in our all-cohort analysis. This cluster also comprised of the most anteriorly-located subregion of the corpus callosum, known as the genu. Adult findings also demonstrated significantly increased volume of the paracentral lobule, as well as slightly increased volume in the right precuneus which was decreased in the entire cohort.

The genu is associated with flexibility during reward-based learning^133^, and has not been previously reported as abnormal in AN^134,135^. PET studies in humans with AN found significantly increased serotonergic receptor activity within the subgenual cingulate, within the region of the genu^136^. The genu also bridges the gap between the limbs of the internal capsules, which have been associated in AN^137-139^ and have been prime targets for deep-brain stimulation in recovery protocols^140-142^. Volumetric increase seen in the genu or internal capsule may reflect alterations in reward-based learning, and development of attentional biases to disorder-salient stimuli^137^. Age-related changes between adult populations compared with all collated data in AN led to a significant effect in the ACC, widely reported throughout the AN literature. The central location of the anterior cingulate cortex within the brain reflects its functional role as a complex information processing hub, performing roles in cognitive and emotional control, such as the processing of universal emotions, conflict monitoring, appraisal of emotional stimuli, error prediction, modulation of risk/fear-mediated behaviour and conditioning^143-149^.

Contrasting our findings, previous work into the ACC in those with AN show reduction in volume^134,150,151^ and inconsistent recovery success of ACC thickness following treatment protocols^70^. fMRI studies in AN find increased activity of the ACC, particularly during set-shifting tasks assessing cognitive flexibility, which was associated with traits of perfectionism, elevated monitoring and intolerance of uncertainty^152^. The ACC is also densely connected to the paracentral lobules which together play important roles in functional integration^153^. Development of the ACC appears to be particularly important during adolescence, and participates in significant structural reorganization^154,155^. Transformation of the ACC during adolescence may provide an explanation as to why no ACC clusters were visualised when testing the entire AN cohort which included adolescent studies. Regarding age-related OFC findings, structural abnormality is not as pronounced when comparing adults with AN to the entire dataset. It may be possible that adolescents experience bilateral alteration of the OFC while adults with AN experience alteration of the right OFC.

In summary, those with AN exhibit significant structural increase in executive and emotional brain regions, as well as decreases more posteriorly located and implicated in basic visual/somatosensation integration and cognition. Following the logic that increased volume is attributed to increased activity/usage and vice versa, those with AN may be susceptible to increased monitoring and vigilant behaviour as well as working memory and visual impairments. Increase of the parahippocampal gyrus in this study is a particularly robust finding, and may reflect impaired or overgeneralised recall of autobiographical memories. Reduction of volume in regions pertinent to somatosensation and inhibition control may contribute to pathologically significant conflict avoidance and decreased empathy seen in AN^156,157^. A lack of consistently reported anatomical regions across literature in those with ASD renders this review unable to confirm any overlapping differences in structure. Aside from VBM-related findings, a lack of any discernable shared neuroanatomical aberrance between both conditions is likely due to the symptomatic and diagnostic heterogeneity presented by ASD, as well as condition-related increase in prevalence for a variety of psychiatric comorbidities.

## LIMITATIONS

Certain limitations must be considered alongside the results of this study. Conducting meta-analyses on a large scale increases the risk of methodological heterogeneity interfering with validity of findings. Individual studies utilised different scanning parameters and statistical threshold specifications, which are difficult to standardize in preparation for analysis. Additionally, we did not examine how variation in anorexic duration of illness may have affected results.

Additionally, we included patients who recovered from AN (RAN) as an additional clinical subgroup. Regardless of excluding the majority of RAN patients, there were approximately 6% of RAN patients that we were unable to remove from data. We confined our analysis to focus on grey matter changes due to limited VBM literature on white matter available.

Lastly, we are unable determine the direction of causation between aberrant volume and restrictive eating/chronic starvation seen in AN due to cross-sectional analytic methodology. As is often the case with neuroimaging studies, findings within this study do not elaborate on the causal relationship between unusual neuroanatomy and condition. While we suggest that increased volume of the ACC may further contribute to symptoms of AN, we are unable to confirm whether restriction associated with AN instead contributes to increased ACC volume.

## CONCLUSION

This study presents the largest to-date meta-analysis of collated MRI coordinate literature in AN and ASD populations. Aggregating all available VBM studies of both conditions demonstrates significantly altered neuroanatomy in those with AN when compared with healthy individuals, but no significant effects in those with ASD, and thus no condition-related structural overlap. Lack of significant findings in ASD contrasts a number of previous associated regions in studies, and suggests an inconsistency in the neuroanatomy associated with ASD. Clusters of increased volume in AN occurred in regions attributed to executive function, decision-making and conflict monitoring, as well as consolidation of episodic and autobiographical memories (MFG/OFC/MTL). Adult-only AN studies noted a significant effect in the ACC and genu, which may reflect impaired conflict monitoring, flexibility, reward-based learning and processing of emotional stimuli. This meta-analysis provides updated information pertaining to neuroanatomical structure associated with AN and ASD, as well as novel findings concerning associated areas of increased volume in AN. Our findings corroborate extensive evidence that psychophysiological symptoms exhibited in with AN are reflected in neuroanatomical structure, and brain regions implicated in this review may serve as effective benchmarks for the detection, diagnosis and treatment of AN.

## Data Availability

All data produced in this study are available upon reasonable request to the corresponding author.

## HIGHLIGHTS

- Findings related to neuroanatomical structure in AN/ASD demonstrate overlap and require revisiting
- Meta-analytic findings show structural increase/decrease vs. healthy controls (LPFC/MTL/OFC) in AN, but no clusters found in ASD
- The neuroanatomy associated with ASD is inconsistent, but findings in AN reflect condition-related impairment in executive function and sociocognitive behaviours.

## ACKNOWLEDGEMENTS

I would like to express my thanks and sincere gratitude to the Northwood Charitable Trust for funding my PhD studentship and subsequent research. I would also like to extend my thanks to Dr Kate Tchanturia for her preparation of this special edition of the European Eating Disorder Review focusing on ASD and eating disorders.

