## Supplementary Material for "A Meta-Analytic Investigation of Grey Matter Differences in Anorexia Nervosa and Autism Spectrum Disorder"

***3.0 – RESULTS***

*Supplementary Table 1 (All coordinates used for AN and ASD meta-analysis)*

| **ASD MRI** | **GM Increase** | **Region** | **MNI/TAL Coordinates** | | | **Cluster Extent** | **Z/T/p value** |
| --- | --- | --- | --- | --- | --- | --- | --- |
|  | *0=decrease, 1=increase* |  | *x* | *y* | *z* | **(mm^3)/k** |  |
| *Cai et al, 2018* | 1 | L Inferior Temporal Gyrus | -48 | 1.5 | -42 | 189 | T=4.38 |
|  | 1 | R Middle Temporal Gyrus | 36 | 9 | -42 | 67 | T=3.81 |
|  | 1 | L Middle Temporal Gyrus | -57 | 3 | -21 | 283 | T=4.16 |
|  | 0 | R Cerebellum Anterior Lobe | 18 | -28.5 | -19.5 | 84 | T=-3.93 |
|  | 0 | R Precuneus | 3 | -54 | 60 | 377 | T=-4.63 |
|  | 0 | R Angular | 49.5 | -64.5 | 45 | 167 | T=-4.25 |
| *Kojima et al, 2019* | 0 | Bilateral Posterior Cingulate Gyrus | -10 | -54 | 24 | 6994 k | T= -4.23 |
| *Ni et al, 2018* | 0 | Lateral Occipital Cortex, Superior | -33 | -63 | 51 | 1301 voxels | T= -4.85 |
|  | 0 | Lateral Occipital Cortex, Superior | -29 | -75 | 48 | ^ | T= -4.60 |
|  | 0 | Lateral Occipital Cortex, Superior | -27 | -61 | 44 | ^ | T= -4.57 |
|  | 0 | L Anterior Prefrontal Pole | -7 | 60 | -24 | 909 voxels | T= -4.40 |
|  | 0 | L Anterior Prefrontal Pole | -6 | 60 | -11 | ^ | T= -3.54 |
|  | 0 | L Anterior Prefrontal Pole | -24 | 60 | -11 | ^ | T= -3.50 |
| *Wang et al, 2017* | 1 | L Superior Temporal Gyrus | -50.5 | -9.5 | -7.5 | 2,981 voxels | T=3.23-4.60 |
|  | 1 | Posterior Cingulate Cortex (PCG) | -61.5 | -7.5 | 31.5 | ^ | ^ |
| *Sato et al, 2017* | 0 | R Inferior Occipital Gyrus | 50 | -75 | -3 | 15,623 | Z= -4.12 |
|  | 0 | R Inferior Temporal Gyrus | 53 | -51 | -11 | ^ | Z= -3.89 |
|  | 0 | R Middle Temporal Gyrus | 71 | -41 | -9 | ^ | Z= -3.48 |
|  | 0 | R Middle Temporal Gyrus | 66 | -41 | 3 | ^ | Z= -3.41 |
|  | 0 | R Middle Temporal Gyrus | 69 | -29 | -3 | ^ | Z= -3.23 |
|  | 0 | L Fusiform Gyrus | -33 | -50 | -5 | 10,702 | Z= -3.19 |
|  | 0 | L Amygdala | -18 | 3 | -14 | ^ | Z= -3.21 |
|  | 0 | R Amygdala | 29 | -12 | -32 | 10,800 | Z= -3.50 |
|  | 0 | R Parahippocampal Gyrus | 33 | -11 | -29 | ^ | Z= -3.20 |
|  | 0 | R Middle Frontal Gyrus | 36 | 27 | 53 | 7,830 | Z= -3.31 |
|  | 0 | R Middle Frontal Gyrus | 27 | 53 | 32 | ^ | Z= -3.70 |
|  | 0 | R Supplementary Motor Area | 11 | 6 | 48 | 21,769 | Z= -3.79 |
|  | 0 | L Middle Cingulate Gyrus | -11 | 9 | 42 | ^ | Z= -4.50 |
|  | 0 | R Middle Cingulate Gyrus | 12 | 18 | 41 | ^ | Z= -3.78 |
|  | 0 | L Medial Superior Frontal Gyrus | -3 | 33 | 50 | ^ | Z= -3.77 |
|  | 0 | L Anterior Cingulate Gyrus | -12 | 33 | 29 | ^ | Z= -2.89 |
|  | 0 | L Middle Frontal Gyrus | -20 | 35 | 44 | ^ | Z= -2.64 |
|  | 0 | L Medial Superior Frontal Gyrus | -8 | 41 | 38 | ^ | Z= -3.92 |
|  | 0 | R Medial Orbital Gyrus | -23 | 35 | 44 | ^ | Z= -3.15 |
| *Parellada et al, 2017* | 0 | Posterior Insula | -40 | -11 | 1 | 31 voxels | p= 0.0098 |
|  | 0 | Posterior Insula | -42 | -5 | -7 | 335 voxels | p= 0.012 |
| *Yang et al, 2016* | 1 | R Inferior Parietal | 38.9 | -78.1 | 20.8 | 2275.23 mm^2 | T= 3.862 |
|  | 1 | R Inferior Temporal | 51.2 | -13.5 | -30.5 | 1625.12 mm^2 | T= 3.820 |
|  | 1 | L Isthmus Cingulate | -13.5 | -48.8 | 6.2 | 4186.02 mm^2 | T= 3.645 |
|  | 1 | R Lingual | 10.4 | -91.7 | -1.7 | 1738.54 mm^2 | T= 3.644 |
| *Hanaie et al, 2016* | 0 | Brainstem | 5 | -40 | -35 | 85 voxels | Z= 3.35 |
|  | 0 | L Intraparietal Sulcus | -35 | -45 | 28 | 63 voxels | Z= 2.84 |
| *Retico et al, 2016* | 1 | L Superior Temporal Gyrus | -49 | -19 | -6 | 114 voxels | p<0.001 |
|  | 1 | L Superior Frontal Gyrus | -25 | 47 | 13 | 267 voxels | p<0.001 |
|  | 1 | R Superior Temporal Gyrus | 44 | -37 | 7 | 128 voxels | p<0.001 |
|  | 1 | R Superior Frontal Gyrus | 25 | 44 | 18 | 74 voxels | p<0.001 |
|  | 1 | L Middle Occipital Gyrus | -26 | -80 | 12 | 57 voxels | p<0.001 |
|  | 1 | L Precuneus | -25 | -68 | 26 | 128 voxels | p<0.001 |
|  | 1 | R Precuneus | 5 | -50 | 29 | 152 voxels | p<0.001 |
|  | 1 | L Precuneus | -7 | -47 | 30 | 182 voxels | p<0.001 |
|  | 0 | R Inferior Temporal Gyrus | 45 | -50 | 4 | 83 voxels | p<0.001 |
| *Floris et al, 2016* | 1 | R Inferior Parietal Lobule | 45 | -54 | 22 | 431 voxels | T= 4.61 |
| *Lin et al, 2015* | 1 | L Medial Dorsal Nucleus of Thalamus | -1 | -12 | 19 | 563 voxels | T= 4.71 |
|  | 1 | L Medial Dorsal Nucleus of Thalamus | 0 | -6 | 9 | ^ | T= 4.09 |
|  | 1 | R Lateral Dorsal Nucleus of Thalamus | 11 | -21 | 15 | ^ | T= 3.71 |
|  | 1 | R Inferior Frontal Gyrus | 3 | 8 | -24 | 761 voxels | T= 4.68 |
|  | 1 | R Inferior Frontal Gyrus | 27 | 15 | -23 | ^ | T= 4.37 |
|  | 1 | R Inferior Frontal Gyrus | 30 | 27 | -27 | ^ | T= 4.17 |
|  | 0 | L Angular/Middle Occipital Gyrus | -40 | -75 | 30 | 430 voxels | T= 3.81 |
|  | 0 | L Precuneus/Middle Occipital Gyrus | -27 | -84 | 40 | ^ | T= 3.75 |
|  | 0 | L Precuneus/Middle Occipital Gyrus | -36 | -75 | 42 | ^ | T= 3.51 |
|  | 1 | L Medial Dorsal Nucleus of Thalamus | -1 | -12 | 19 | 697 voxels | T= 4.79 |
|  | 1 | R Lateral Dorsal Nucleus of Thalamus | 11 | -21 | 15 | ^ | T= 3.93 |
|  | 1 | B Lateral Dorsal Nucleus of Thalamus | 0 | 3 | 18 | ^ | T= 3.43 |
|  | 1 | R Inferior Frontal Gyrus | 3 | 8 | -24 | 605 voxels | T= 4.63 |
|  | 1 | R Inferior Frontal Gyrus | 27 | 15 | -3 | ^ | T= 4.35 |
|  | 1 | R Inferior Frontal Gyrus | 30 | 27 | -27 | ^ | T= 4.10 |
| *Lim et al, 2015* | 1 | L Middle/Superior Temporal Gyrus | -50 | 3 | -17 | 1956 voxels | p= 0.04 |
|  | 1 | L Medial Frontal Gyrus | -12 | -9 | 57 | 516 voxels | p= 0.049 |
| *Radeloff et al, 2014* | 0 | L Amygdala | -17 | -5 | -15 | 319 voxels | Z= 4.10 |
|  | 0 | R Amygdala | 27 | -9 | -18 | 39 voxels | Z= 3.37 |
|  | 0 | L Medial Prefrontal Cortex | -2 | 48 | 15 | 28 voxels | Z= 3.56 |
|  | 0 | L Insula | -30 | 15 | 18 | 22 voxels | Z= 3.54 |
|  | 1 | R Occipital Medial Area | 39 | -75 | 23 | 118 voxels | Z= 3.95 |
| *Xiao et al, 2014* | 1 | R Superior Temporal Gyrus | 52 | -10 | 5 | 66 voxels | T= 3.50 |
|  | 1 | R Temporal Lobe | 24 | 9 | -42 | 50 voxels | T= 3.38 |
|  | 1 | R Superior Temporal Gyrus | 56 | -12 | 0 | 260 voxels | T= 4.18 |
|  | 1 | L Middle Temporal Gyrus | -44 | 8 | -32 | 88 voxels | T= 3.97 |
|  | 1 | R Insula | 47 | -15 | 5 | 73 voxels | T= 3.75 |
|  | 1 | R Heschl | 35 | -19 | 9 | 63 voxels | T= 3.47 |
| *Ecker et al, 2013* | 1 | L Superior Frontal | -19.7 | 0.99 | 57.33 | N/A | T= 2.938 |
|  | 1 | L Superior Frontal | -9.62 | 13.86 | 43.58 | N/A | T= 2.749 |
|  | 1 | L Superior Frontal | -15.95 | 43.83 | 29.03 | N/A | T= 3.295 |
|  | 1 | L Caudal Middle Frontal | -38.19 | 5.92 | 43.43 | N/A | T= 2.398 |
|  | 1 | R Caudal Middle Frontal | 35.93 | 7.77 | 42.99 | N/A | T= 2.348 |
|  | 1 | L Rostral Middle Frontal | -35.64 | 20.46 | 22.51 | N/A | T= 2.983 |
|  | 1 | L Rostral Middle Frontal | -35.6 | 42.5 | 3.78 | N/A | T= 3.095 |
|  | 1 | R Rostral Middle Frontal | 34.69 | 39.93 | 13.87 | N/A | T= 2.606 |
|  | 1 | L Pars Opercularis | -41.83 | 17.66 | 10.75 | N/A | T= 2.601 |
|  | 1 | R Pars Triangularis | 45.45 | 21.88 | 17.51 | N/A | T= 2.056 |
|  | 1 | R Medial Orbitofrontal | 8.5 | 48.45 | 4.04 | N/A | T= 2.658 |
|  | 1 | R Middle Temporal Gyrus | 55.89 | -44.54 | 3.11 | N/A | T= 3.219 |
|  | 1 | R Superior Temporal Gyrus | 52.94 | -33.07 | 14.73 | N/A | T= 3.182 |
|  | 1 | L Inferior Parietal | -40.78 | -69.6 | 26.7 | N/A | T= 2.635 |
|  | 1 | R Supramarginal Gyrus | 33.53 | -32.68 | 44.06 | N/A | T= 2.285 |
|  | 1 | L Supramarginal Gyrus | -47.83 | -24.44 | 28.97 | N/A | T= 2.743 |
|  | 1 | R Superior Parietal | 22.89 | -57.48 | 28.77 | N/A | T= 2.092 |
|  | 1 | L Lateral Occipital Cortex | -37.91 | -78.33 | 2.3 | N/A | T= 2.313 |
|  | 1 | R Lateral Occipital Cortex | 42.92 | -72.33 | 8.68 | N/A | T= 2.562 |
|  | 1 | R Postcentral Gyrus | 35.52 | -20.97 | 33.55 | N/A | T= 3.309 |
|  | 1 | L Postcentral Gyrus | -54.03 | -11.13 | 13.36 | N/A | T= 3.147 |
|  | 1 | L Posterior Cingulate | -3.79 | -10.03 | 27.57 | N/A | T= 2.393 |
|  | 0 | L Anterior Cingulate | -5.27 | 8.67 | -1.19 | N/A | T= -2.349 |
|  | 0 | R Medial Prefrontal Cortex | 9.57 | 30.44 | -9.57 | N/A | T= -2.244 |
|  | 0 | R Medial Prefrontal Cortex | 17.94 | -11.19 | 49.66 | N/A | T= -2.477 |
|  | 0 | L Middle Temporal Gyrus | -55.3 | -8.58 | -14.43 | N/A | T= -3.003 |
|  | 0 | R Middle Temporal Gyrus | 53.24 | -26.77 | -9.31 | N/A | T= -2.912 |
|  | 0 | R Inferior Temporal Gyrus | 49.73 | -16.06 | -22.21 | N/A | T= -3.060 |
|  | 0 | L Parahippocampal Gyrus | -23.37 | -12.27 | -19.14 | N/A | T= -2.331 |
|  | 0 | L Superior Parietal | -17.92 | -69.22 | 35.48 | N/A | T= 2.387 |
|  | 0 | R Superior Parietal | 5.6 | -49.47 | 54.2 | N/A | T= 2.133 |
|  | 0 | L Supramarginal gyrus | -49.55 | -37.63 | 39 | N/A | T= -2.227 |
|  | 0 | L Precuneus | -8.81 | -46.31 | 42.66 | N/A | T= -2.409 |
|  | 0 | R Pericalcarine fissure | 12.89 | -78.77 | 3.94 | N/A | T= -3.578 |
|  | 0 | R Lingual Gyrus | 17.48 | -55.98 | 2.47 | N/A | T= -2.883 |
| *Greimel et al, 2013* | 0 | B Anterior Cingulate Cortex | 0 | 42 | 12 | > 100 voxels | N/A |
|  | 0 | R Posterior Superior Temporal Gyrus | 68 | -34 | -44 | > 100 voxels | N/A |
|  | 0 | R Posterior Superior Temporal Gyrus | 62 | -30 | -12 | > 100 voxels | N/A |
| *Mengotti et al, 2011* | 1 | R Inferior Parietal Cortex | 52 | -57 | 29 | 1253 voxels | T= 4.87 |
|  | 1 | R Inferior Parietal Cortex | 44 | -68 | 35 | 175 voxels | T= 3.90 |
|  | 1 | R Superior Occipital Gyrus | 23 | -88 | 12 | 245 voxels | T= 4.01 |
|  | 1 | L Inferior Temporal Gyrus | -52 | -55 | -10 | 417 voxels | T= 4.38 |
|  | 1 | R Inferior Temporal Gyrus | 56 | -52 | -18 | 236 voxels | T= 4.02 |
|  | 1 | L Superior Parietal Lobule | -36 | -50 | 54 | 210 voxels | T= 4.10 |
|  | 1 | L Precuneus | -12 | -45 | 73 | 127 voxels | T= 3.66 |
|  | 0 | R Inferior Frontal Gyrus | 37 | 28 | 27 | 131 voxels | T= 4.03 |
|  | 0 | L Supplementary Motor Area | -11 | -10 | 50 | 315 voxels | T- 4.97 |
| *Toal et al, 2010* | 0 | R Inferior Temporal Gyrus | 30 | -4.9 | -31.6 | 896 voxels | p= 0.003 |
|  | 0 | L Parahippocampal Gyrus | -25.1 | -25.3 | -24.8 | 818 voxels | p= 0.003 |
|  | 1 | R Superior Temporal Gyrus | 55.9 | -28.1 | 19.3 | 1227 voxels | p= 0.003 |
|  | 1 | R Cingulate Gyrus | 0.5 | -31 | 43.3 | 1148 voxels | p= 0.003 |
|  | 0 | L Brainstem | -2.4 | -25 | -29.7 | 1008 voxels | p= 0.01 |
|  | 0 | R Cerebrum/Corpus Callosum | 2.2 | 25 | 9.6 | 584 voxels | p= 0.01 |
| *Wilson et al, 2009* | 0 | L Cerebellar Lobule VIIB | -44 | -44 | -47 | 711 voxels | T= 4.36 |
|  | 0 | L Cerebellar Lobule VIII | -30 | -30 | -52 | 82 voxels | T= 3.90 |
|  | 0 | R Cerebellar Crus I | 55 | 55 | -37 | 204 voxels | T= 4.16 |
| *Hyde et al, 2010* | 1 | R Brainstem/midbrain | 14 | -11 | -4 | N/A | T= 4.243 |
|  | 1 | L Brainstem/midbrain | -10 | -12 | -7 | N/A | T= 3.76 |
|  | 1 | L Brainstem/Reticular Formation | -7 | -41 | -32 | N/A | T= 4.251 |
|  | 1 | R Medial Frontal Gyrus | 6 | 69 | -12 | N/A | T= 3.911 |
|  | 1 | L Medial Orbital Frontal Gyrus | -9 | 44 | -11 | N/A | T= 3.716 |
|  | 1 | R Middle Frontal Gyrus | 28 | 50 | 16 | N/A | T= 5.566 |
|  | 1 | R Middle Frontal Gyrus | 26 | 42 | 12 | N/A | T= 3.967 |
|  | 1 | L Middle Frontal Gyrus | -34 | 43 | 13 | N/A | T= 3.958 |
|  | 0 | R Postcentral Gyrus | 40 | -33 | 57 | N/A | T= -4.264 |
|  | 0 | R Precentral Gyrus | 30 | -23 | 65 | N/A | T= -3.956 |
|  | 0 | L Precentral Gyrus | -19 | -21 | 69 | N/A | T= -3.94 |
|  | 0 | L Anterior Cerebellum | -2 | -60 | -6 | N/A | T= -5.713 |
|  | 0 | R Brainstem/midbrain | 14 | -12 | -4 | N/A | T= -4.22 |
|  | 0 | R Brainstem/midbrain | 1 | -20 | -13 | N/A | T= -4.062 |
| *Brieber et al, 2007* | 1 | R Supramarginal Gyrus | 47 | -39 | 29 | 50 voxels | Z= 3.71 |
|  | 1 | L Inferior Parietal/Postcentral Gyrus | -31 | -38 | 51 | 65 voxels | Z= 3.37 |
|  | 0 | L Hippocampus-Amygdala | -31 | -7 | -17 | 18 voxels | Z= 3.16 |
| *Rojas et al, 2006* | 1 | L Medial Frontal Gyrus | -8 | 41 | 54 | 1459 voxels | T= 3.43 |
|  | 1 | R Medial Frontal Gyrus | 15 | 58 | 32 | 160 voxels | T= 3.08 |
|  | 1 | L Precentral Gyrus | -37 | -15 | 60 | 693 voxels | T= 3.11 |
|  | 1 | L Middle Temporal Gyrus | -50 | -72 | 21 | 1166 voxels | T= 3.24 |
|  | 1 | R Fusiform Gyrus | 38 | -25 | -28 | 354 voxels | T= 3.095 |
|  | 1 | R Postcentral Gyrus | 50 | -9 | 27 | 1155 voxels | T= 3.76 |
|  | 1 | L Caudate Nucleus | -13 | 21 | -4 | 344 voxels | T= 2.95 |
|  | 1 | R Caudate Nucleus | 9 | 20 | -3 | 179 voxels | T= 2.80 |
|  | 1 | L Hippocampus | -22 | -28 | -6 | 274 voxels | T= 3.48 |
|  | 0 | L Cerebellar Crus I | -46 | -42 | -31 | 11342 voxels | T= 3.33 |
|  | 0 | L Cerebellar Lobule VIII | -25 | -48 | -48 | 299 voxels | T= 2.77 |
|  | 0 | L Cerebellar Lobule IX | -5 | -59 | -59 | 11342 voxels | T= 3.36 |
|  | 0 | R Cerebellar Crus I | 53 | -54 | -36 | 2792 voxels | T= 3.38 |
| *Boddaert et al, 2004* | 0 | R Superior Temporal Sulcus | 38 | -24 | -4 | 440 voxels | Z= 3.75 |
|  | 0 | R Superior Temporal Sulcus | 40 | -34 | -6 | ^ | Z= 3.49 |
|  | 0 | R Superior Temporal Sulcus | 51 | -36 | -9 | ^ | Z= 3.15 |
|  | 0 | L Superior Temporal Sulcus | -38 | -27 | -3 | 195 voxels | Z= 3.59 |
|  | 0 | R Temporal Pole | 50 | 8 | -28 | 814 voxels | Z= 4.90 |
|  | 0 | L Cerebellum | -24 | -60 | -39 | 1209 voxels | Z= 4.99 |
| *Mueller et al, 2013* | 0 | Medial Temporal Gyrus/Inferior Temporal Gyrus | -48 | -22 | -2 | 148 voxels | p= 0.072 |
|  | 0 | R Superior Parietal Lobe/Supramarginal Gyrus/Angular Gyrus/Temporal Lobe/Lateral Occipital Lobe/Temporo-parietal Junction | 44 | -42 | 34 | 540voxels | p<0.0001 |
|  | 0 | R Superior Parietal Lobe/Supramarginal Gyrus/Angular Gyrus/Temporal Lobe/Lateral Occipital Lobe/Temporo-parietal Junction | 22 | -60 | 52 | 145 voxels | p<0.0001 |
|  | 0 | L Medial Temporal Gyrus/Inferior Temporal Gyrus | -64 | -38 | -36 | 1016 voxels | p<0.0001 |
|  | 0 | R Superior Parietal Lobe/Supramarginal Gyrus/Angular Gyrus/Temporal Lobe/Lateral Occipital Lobe/Temporo-parietal Junction | 44 | -42 | 34 | 4392 voxels | p< 0.001 |
|  | 0 | L Superior Parietal Lobe/Supramarginal Gyrus/Angular Gyrus/Temporal Lobe/Lateral Occipital Lobe/Temporo-parietal Junction/ | -46 | -48 | 6 | 6219 voxels | p< 0.001 |
|  | 0 | L Medial Temporal Gyrus/Inferior Temporal Gyrus | -64 | -38 | -26 | 5271 voxels | p< 0.001 |
|  | 0 | R Medial Temporal Gyrus/Inferior Temporal Gyrus | 70 | -24 | -6 | 808 voxels | p= 0.001 |
|  | 0 | R Inferior Frontal Gyrus/Orbito-Frontal Cortex/Frontal Pole | 36 | 40 | 40 | 2472 voxels | p< 0.001 |
|  | 0 | L Inferior Frontal Gyrus/Orbito-Frontal Cortex/Frontal Pole | -50 | 46 | 12 | 826 voxels | p= 0.001 |
|  | 0 | L Frontal Pole | -12 | 38 | -30 | 534 voxels | p< 0.001 |
|  | 0 | L Middle Frontal Gyrus | -30 | 16 | 28 | 202 voxels | p= 0.003 |
|  | 0 | B Medial Prefrontal Cortex/Cingulate Gyrus | -2 | 66 | 28 | 1720 voxels | p< 0.001 |
|  | 0 | L Insula | -42 | -2 | 20 | 164 voxels | p= 0.001 |
| *McAlonan et al, 2009* | 0 | L Frontal Lobe | -18.9 | 1.1 | 16.4 | 1102 voxels | NS |
|  | 1 | L Basal Ganglia | -23.2 | 6 | 4.8 | 1060 voxels | NS |
|  | 1 | R Basal Ganglia | 7.5 | -3.1 | 1.1 | 284 voxels | NS |
| *Duerden et al, 2014* | 1 | R Paracentral Lobule | 78 | -7 | 62.64 | N/A | T= 3.52 |
|  | 1 | L Middle Frontal Gyrus | -31 | 21 | 47 | N/A | T= 3.2 |
| *Bonilha et al, 2008* | 1 | L Inferior Frontal Gyrus | -17 | 29 | -12 | N/A | T= 6.25 |
|  | 1 | L Cuneus | -29 | -75 | 5 | N/A | T= 6.06 |
|  | 1 | L Cingulate Gyrus | -17 | -49 | 21 | N/A | T= 5.82 |
|  | 1 | L Claustrum | -25 | -11 | 17 | N/A | T= 5.72 |
|  | 1 | L Precuneus | 21 | -49 | 43 | N/A | T= 5.72 |
|  | 1 | L Middle Temporal Gyrus | -56 | -34 | -7 | N/A | T= 5.67 |
|  | 1 | L Middle Temporal Gyrus | -37 | -77 | -19 | N/A | T= 5.58 |
|  | 1 | L Thalamus | -5 | -29 | 17 | N/A | T= 5.53 |
|  | 1 | L Superior Frontal Gyrus | -14 | 51 | 28 | N/A | T= 5.49 |
|  | 1 | L Cingulate Gyrus | -17 | 6 | 41 | N/A | T= 5.47 |
|  | 1 | L Superior Parietal Lobule | -25 | -51 | 43 | N/A | T= 5.47 |
|  | 1 | L Inferior Temporal Gyrus | -47 | -26 | -12 | N/A | T= 5.41 |
|  | 1 | L Insula | -41 | -42 | 24 | N/A | T= 5.36 |
|  | 1 | L Putamen | -20 | -5 | 13 | N/A | T= 5.36 |
|  | 1 | L Superior Temporal Gyrus | 50 | -7 | -4 | N/A | T= 5.32 |
|  | 1 | L Anterior Cingulate | 16 | 30 | -9 | N/A | T= 4.98 |
|  | 1 | L Fusiform Gyrus | 37 | -73 | -12 | N/A | T= 4.97 |
|  | 1 | L Superior Frontal Gyrus | -11 | 36 | 45 | N/A | T= 4.94 |
|  | 1 | L Middle Occipital Gyrus | -20 | -91 | 8 | N/A | T= 4.93 |
|  | 1 | L Cuneus | -18 | -87 | 21 | N/A | T= 4.87 |
|  | 1 | L Lingual Gyrus | -23 | -79 | 3 | N/A | T= 4.83 |
|  | 1 | L Precentral Gyrus | -37 | -8 | 26 | N/A | T= 4.71 |
|  | 1 | L Superior Temporal Gyrus | -39 | -52 | 32 | N/A | T= 4.71 |
|  | 1 | L Middle Temporal Gyrus | 50 | -55 | -2 | N/A | T= 4.69 |
|  | 1 | L Postcentral Gyrus | -25 | -44 | 61 | N/A | T= 4.67 |
|  | 1 | L Cerebellar Declive | 36 | -83 | -18 | N/A | T= 4.6 |
|  | 1 | L Inferior Occipital Gyrus | -33 | -84 | -9 | N/A | T= 4.58 |
|  | 1 | L Cerebellar Pyramis | 30 | -76 | -31 | N/A | T= 4.46 |
|  | 1 | L Inferior Frontal Gyrus | 33 | 31 | 10 | N/A | T= 4.45 |
|  | 1 | L Superior Frontal Gyrus | -16 | 59 | 9 | N/A | T= 4.42 |
|  | 1 | L Cingulate Gyrus | -15 | -2 | 47 | N/A | T= 4.41 |
|  | 1 | L Superior Frontal Gyrus | -15 | 9 | 58 | N/A | T= 4.38 |
|  | 1 | L Middle Temporal Gyrus | -38 | -53 | -5 | N/A | T= 4.36 |
|  | 1 | L Thalamus | -27 | -28 | 6 | N/A | T= 4.34 |
|  | 1 | L Fusiform Gyrus | 48 | -41 | -14 | N/A | T= 4.33 |
|  | 1 | L Parahippocampal Gyrus | 17 | -48 | -3 | N/A | T= 4.3 |
|  | 1 | L Medial Frontal Gyrus | 5 | 37 | -15 | N/A | T= 4.28 |
|  | 1 | L Inferior Frontal Gyrus | -45 | 33 | 4 | N/A | T= 4.27 |
|  | 1 | L Medial Frontal Gyrus | -19 | 34 | 28 | N/A | T= 4.28 |
|  | 1 | R Superior Frontal Gyrus | 10 | 36 | 44 | N/A | T= 5.97 |
|  | 1 | R Inferior Occipital Gyrus | 23 | -90 | -7 | N/A | T= 5.86 |
|  | 1 | R Precuneus | 21 | -49 | 43 | N/A | T= 5.72 |
|  | 1 | R Cingulate Gyrus | 23 | -19 | 39 | N/A | T= 5.34 |
|  | 1 | R Superior Temporal Gyrus | 50 | -7 | -4 | N/A | T= 5.32 |
|  | 1 | R Superior Frontal Gyrus | 19 | 56 | -7 | N/A | T= 5.27 |
|  | 1 | R Cingulate Gyrus | 16 | -48 | 21 | N/A | T= 5.27 |
|  | 1 | R Posterior Cingulate | 4 | -27 | 19 | N/A | T= 5.01 |
|  | 1 | R Anterior Cingulate | 16 | 30 | -9 | N/A | T= 4.98 |
|  | 1 | R Fusiform Gyrus | 37 | -73 | -12 | N/A | T= 4.97 |
|  | 1 | R Superior Parietal Lobule | 17 | -53 | 58 | N/A | T= 4.87 |
|  | 1 | R Superior Frontal Gyrus | 20 | 8 | 56 | N/A | T= 4.86 |
|  | 1 | R Medial Frontal Gyrus | 18 | 50 | 6 | N/A | T= 4.84 |
|  | 1 | R Thalamus | 26 | -28 | 6 | N/A | T= 4.81 |
|  | 1 | R Cuneus | 16 | -80 | 17 | N/A | T= 4.75 |
|  | 1 | R Middle Temporal Gyrus | 50 | -55 | -2 | N/A | T= 4.69 |
|  | 1 | R Cerebellar Declive | 36 | -83 | -18 | N/A | T= 4.6 |
|  | 1 | R Thalamus | 24 | -21 | 14 | N/A | T= 4.52 |
|  | 1 | R Cerebellar Pyramis | 30 | -76 | -31 | N/A | T= 4.46 |
|  | 1 | R Inferior Frontal Gyrus | 33 | 31 | 10 | N/A | T= 4.45 |
|  | 1 | R Insula | 32 | -7 | 20 | N/A | T= 4.39 |
|  | 1 | R Middle Frontal Gyrus | 20 | 1 | 60 | N/A | T= 4.36 |
|  | 1 | R Superior Temporal Gyrus | 41 | -49 | 25 | N/A | T= 4.34 |
|  | 1 | R Fusiform Gyrus | 48 | -41 | -14 | N/A | T= 4.33 |
|  | 1 | R Parahippocampal Gyrus | 17 | -46 | -3 | N/A | T= 4.3 |
|  | 1 | R Medial Frontal Gyrus | 5 | 37 | -15 | N/A | T= 4.28 |
|  | 1 | R Caudate Nucleus | 39 | -24 | -7 | N/A | T= 4.25 |
|  | 1 | R Middle Temporal Gyrus | 43 | -70 | -16 | N/A | T= 4.22 |
|  | 1 | R Medial Frontal Gyrus | 16 | 45 | 17 | N/A | T= 4.16 |
|  | 1 | R Inferior Parietal Lobule | 36 | -59 | 44 | N/A | T= 4.07 |
|  | 0 | L Cuneus | -46 | -13 | -21 | N/A | T= 6.32 |
|  | 0 | L Middle temporal gyrus | 10 | 37 | 44 | N/A | T= 6.22 |
|  | 0 | L Sub-gyral | 12 | 28 | 45 | N/A | T= 6.02 |
|  | 0 | L inferior frontal gyrus | -39 | 39 | -11 | N/A | T= 5.93 |
|  | 0 | L Middle frontal gyrus | -58 | -53 | 25 | N/A | T= 5.72 |
|  | 0 | L Supramarginal gyrus | 17 | 5 | 50 | N/A | T= 5.3 |
|  | 0 | L Parahippocampal gyrus | 17 | 37 | -19 | N/A | T= 4.82 |
|  | 0 | L uncus | -23 | -15 | -36 | N/A | T= 4.45 |
|  | 0 | L Postcentral gyrus | 33 | 32 | 9 | N/A | T= 4.41 |
|  | 0 | L cerebellar tonsil | 53 | -17 | 18 | N/A | T= 4.3 |
|  | 0 | L superior frontal gyrus | 23 | -7 | -33 | N/A | T= 4.14 |
|  | 0 | L Precentral gyrus | -22 | -9 | 71 | N/A | T= 3.99 |
|  | 0 | L cingulate gyrus | -35 | -9 | 47 | N/A | T= 3.820 |
|  | 0 | L superior temporal gyrus | -2 | 63 | 2 | N/A | T= 3.53 |
|  | 0 | L Medial frontal gyrus | 12 | 59 | -24 | N/A | T= 3.49 |
|  | 0 | L precuneus | 18 | 32 | -30 | N/A | T= 3.28 |
|  | 0 | L anterior cingulate | 60 | -45 | 12 | N/A | T= 3.21 |
|  | 0 | L Thalamus | -7 | 57 | -15 | N/A | T= 3.06 |
|  | 0 | L insula | 19 | 28 | -28 | N/A | T= 2.97 |
|  | 0 | L fusiform gyrus | 48 | -37 | 25 | N/A | T= 2.93 |
|  | 0 | R Superior Temporal gyrus | 59 | -18 | -5 | N/A | T= 7.28 |
|  | 0 | R superior frontal gyrus | 12 | 36 | 55 | N/A | T= 6.07 |
|  | 0 | R middle frontal gyrus | 14 | -1 | 66 | N/A | T= 5.19 |
|  | 0 | R superior parietal | 37 | -54 | 3 | N/A | T= 5.18 |
|  | 0 | R cuneus | 44 | -70 | 16 | N/A | T= 5.02 |
|  | 0 | R inferior temporal gyrus | 32 | -75 | 4 | N/A | T= 4.84 |
|  | 0 | R anterior cingulate | 8 | 26 | -8 | N/A | T= 4.68 |
|  | 0 | R medial frontal gyrus | -17 | 30 | -12 | N/A | T= 4.4 |
|  | 0 | R postcentral gyrus | 51 | -57 | -2 | N/A | T= 4.36 |
|  | 0 | R Middle temporal gyrus | -57 | -34 | -7 | N/A | T= 4.35 |
|  | 0 | R cerebellar tonsil | -21 | -35 | -39 | N/A | T= 4.31 |
|  | 0 | R uncus | -16 | -27 | 69 | N/A | T= 4.13 |
|  | 0 | R inferior frontal gyrus | 18 | 50 | 6 | N/A | T= 3.9 |
|  | 0 | R parahippocampal gyrus | 42 | 2 | -35 | N/A | T= 3.9 |
|  | 0 | R insula | 64 | 24 | 0 | N/A | T= 3.57 |
|  | 0 | R posterior cingulate | -52 | 0 | -8 | N/A | T= 3.46 |
|  | 0 | R inferior semi-lunar lobule | -46 | 17 | 50 | N/A | T= 3.38 |
|  | 0 | R precuneus | 4 | 61 | -10 | N/A | T= 3.32 |
|  | 0 | R precentral gyrus | -14 | -63 | 43 | N/A | T= 3.31 |
|  | 0 | R orbital gyrus | -37 | 5 | 46 | N/A | T= 3.24 |
|  | 0 | R inferior parietal lobule | -4 | 9 | 21 | N/A | T= 3.23 |
|  | 0 | R cingulate gyrus | -73 | -8 | 26 | N/A | T= 3.04 |
|  | 0 | R culmen | 26 | 37 | 48 | N/A | T= 3 |
| *Calderoni et al, 2012* | 1 | L Superior Frontal Gyrus | -26 | 44 | 20 | 116 voxels | p< 0.05 |
|  | 1 | R Superior Frontal Gyrus | 26 | 50 | 10 | 34 voxels | p< 0.05 |
|  | 1 | R Temporo-Parietal Junction | 45 | -55 | 26 | 24 voxels | p< 0.05 |
| *Cheng et al, 2011* | 0 | R Inferior Frontal Gyrus | 32 | 30 | -10 | 180 | Z= 3.74 |
|  | 0 | R Precentral Gyrus | 44 | -13 | 44 | 306 | Z= 3.58 |
|  | 0 | L Postcentral Gyrus | -39 | -22 | 54 | 57 | Z= 3.48 |
|  | 0 | L Cuneus | -26 | -88 | 26 | 91 | 3.35 |
|  | 0 | L Thalamus | -8 | -22 | 0 | 35 | Z= 3.31 |
|  | 0 | L Lingual Gyrus | -4 | -96 | -1 | 23 | Z= 3.24 |
|  | 0 | R Superior Temporal Gyrus | 52 | -32 | 6 | 13 | Z= 3.22 |
|  | 1 | L Anterior Cingulate | -8 | 42 | -5 | 478 | Z= 4.36 |
|  | 1 | R Paracentral Lobule | 3 | -43 | 56 | 370 | Z= 4.22 |
|  | 1 | L Superior Parietal Lobule | -13 | -55 | 60 | 195 | Z= 3.81 |
|  | 1 | L Precuneus | -4 | -42 | 52 | 122 | Z= 3.79 |
|  | 1 | R Medial Frontal Gyrus | 7 | 61 | 15 | 59 | Z= 3.72 |
|  | 1 | L Fusiform Gyrus | -51 | -25 | -23 | 34 | Z= 3.47 |
|  | 1 | R Middle Frontal Gyrus | 42 | 19 | 45 | 17 | Z= 3.28 |
|  | 1 | L Subcallosal Gyrus | -5 | 18 | -14 | 33 | Z= 3.28 |
| *Foster et al, 2015* | 1 | R Central Sulcus | 32 | -16 | 44 | N/A | T= 4.45 |
|  | 1 | L Medial Frontal Gyrus | -7 | 56 | 30 | N/A | T= 4.36 |
|  | 1 | L Inferior Frontal Gyrus | -54 | 35 | 0 | N/A | T= 4.30 |
|  | 1 | L Precentral Gyrus | -54 | 5 | 35 | N/A | T= 4.18 |
|  | 1 | R Precentral Sulcus | 34 | -1 | 45 | N/A | T= 4.02 |
|  | 1 | L Middle Frontal Gyrus | -49 | 31 | 28 | N/A | T= 3.99 |
|  | 1 | L Presupplementary Motor Area | -7 | 4 | 51 | N/A | T= 3.76 |
|  | 1 | L Medial Precentral Gyrus | -5 | -20 | 65 | N/A | T= 3.71 |
|  | 1 | R Superior Frontal Sulcus | 27 | 47 | 23 | N/A | T= 3.70 |
|  | 1 | L Middle Frontal Gyrus | -15 | 19 | 35 | N/A | T= 3.66 |
|  | 1 | L Anterior Cingulate | -12 | 36 | 30 | N/A | T= 3.57 |
|  | 1 | R Superior Frontal Gyrus | 13 | 71 | 2 | N/A | T= 3.56 |
|  | 1 | L Orbitofrontal Cortex | -38 | 5 | -14 | N/A | T= 3.54 |
|  | 1 | L Medial Frontal Gyrus | -9 | 63 | 11 | N/A | T= 3.51 |
|  | 1 | L Orbitofrontal Cortex | -45 | 34 | -16 | N/A | T= 3.42 |
|  | 1 | R Inferior Frontal Gyrus | 50 | 36 | 15 | N/A | T= 3.42 |
|  | 1 | L Inferior Temporal Gyrus | -59 | -65 | -12 | N/A | T= 4.67 |
|  | 1 | L Middle Temporal Gyrus | -61 | -55 | 8 | N/A | T= 4.30 |
|  | 1 | R Middle Temporal Gyrus | 56 | -26 | -9 | N/A | T= 4.08 |
|  | 1 | L Heschl's Gyrus | -47 | -12 | 2 | N/A | T= 4.02 |
|  | 1 | L Fusiform Gyrus | -18 | -58 | -2 | N/A | T= 4.01 |
|  | 1 | R Middle Temporal Gyrus | 64 | -18 | -13 | N/A | T= 3.98 |
|  | 1 | L Middle Temporal Gyrus | -65 | -16 | -18 | N/A | T= 3.95 |
|  | 1 | R Lingual Gyrus | 3 | -75 | 0 | N/A | T= 3.76 |
|  | 1 | L Fusiform Gyrus | -31 | -44 | -11 | N/A | T= 3.74 |
|  | 1 | L Inferior Temporal Gyrus | -54 | -68 | 5 | N/A | T= 3.62 |
|  | 1 | R Middle Temporal Gyrus | 59 | -36 | -4 | N/A | T= 3.56 |
|  | 1 | L Superior Temporal Gyrus | -43 | 8 | -20 | N/A | T= 3.54 |
|  | 1 | L Poscentral Gyrus | -53 | -11 | 34 | N/A | T= 4.93 |
|  | 1 | L Posterior Cingulate | -9 | -39 | 41 | N/A | T= 4.30 |
|  | 1 | L Precuneus | -10 | -53 | 61 | N/A | T= 3.99 |
|  | 1 | L Precuneus | -7 | -58 | 25 | N/A | T= 3.79 |
|  | 1 | R Supramarginal Gyrus | 42 | -38 | 42 | N/A | T= 3.79 |
|  | 1 | R Angular Gyrus | 41 | 66 | 41 | N/A | T= 3.59 |
|  | 1 | L inferior Occipital Gyrus | -52 | -75 | -2 | N/A | T= 4.19 |
|  | 1 | L Inferior Occipital Gyrus | -33 | -92 | -10 | N/A | T= 4.11 |
|  | 1 | R Cuneus | 3 | -86 | 18 | N/A | T= 3.99 |
|  | 1 | R Cuneus | 8 | -78 | 9 | N/A | T= 3.49 |
|  | 1 | L Cuneus | -9 | -92 | 22 | N/A | T= 3.43 |
|  | 1 | L Putamen | -27 | 3 | -9 | N/A | T= 4.24 |
|  | 1 | L Caudate Nucleus | -19 | 18 | 9 | N/A | T= 3.79 |
|  | 1 | L Caudate Nucleus | -19 | 11 | 17 | N/A | T= 3.66 |
|  | 0 | R Superior Temporal Gyrus | 63 | -44 | 23 | N/A | T= 3.75 |
|  | 0 | R Supramarginal Gyrus | 67 | -14 | 23 | N/A | T= 4.07 |
|  | 0 | R Angular Gyrus | 64 | -40 | 41 | N/A | T= 3.86 |
|  | 0 | L Supramarginal Gyrus | -55 | -33 | 55 | N/A | T= 3.56 |
|  | 0 | L Cerebellum | -10 | -72 | -50 | N/A | T= 3.48 |
| *Kurth et al, 2011* | 0 | L Hypothalamus | -3 | 1 | 22 | 13 voxels | T= 5.24 |
| *Riva et al, 2013* | 0 | L Cerebellum Crus 2 | -44 | -58 | -42 | 424 voxels | T= 2.71 |
|  | 0 | R Cerebellum Crus 2 | 48 | -57 | -42 | 150 voxels | T= 2.06 |
|  | 0 | R Vermis | 6 | -60 | -35 | 749 voxels | T= 2.21 |
|  | 0 | L Hippocampus | -14 | -4 | -18 | 69 voxels | T= 2.26 |
|  | 0 | R Hippocampus | 18 | -6 | -14 | 20 voxels | T= 2.00 |
|  | 0 | L Inferior Frontal Gyrus Pars Triangularis | -48 | 26 | 6 | 327 voxels | T= 3.02 |
|  | 0 | L Occipital Medial Gyrus | -28 | -91 | 13 | 280 voxels | T= 2.99 |
|  | 0 | L Inferior Temporal Gyrus | 36 | -7 | -35 | 370 voxels | T= 2.82 |
|  | 0 | L Occipital Medial Gyrus | -30 | -82 | 27 | 109 voxels | T= 2.47 |
|  | 0 | L Superior Frontal Gyrus | -3 | 39 | 40 | 55 voxels | T= 2.30 |
|  | 0 | L Occipital Superior Gyrus | -15 | -99 | 19 | 97 voxels | T= 2.24 |
|  | 0 | R Occipital Inferior Gyrus | 32 | -85 | -14 | 165 voxels | T= 3.40 |
|  | 0 | R Postcentral Gyrus | 20 | -43 | 64 | 727 voxels | T= 3.34 |
| *Waiter et al, 2005* | 0 | R extra nuclear | 25 | -5 | 25 | N/A | Z= 4.80 |
| *Craig et al, 2007* | 0 | R cingulate gyrus | 16 | -16 | 32 | N/A | Z= 4.74 |
|  | 0 | R middle frontal gyrus | 31 | 48 | 10 | N/A | Z= 4.36 |
|  | 0 | R supramarginal gyrus | 45 | -56 | 32 | N/A | Z= 3.62 |
|  | 0 | R medial frontal gyrus | 6 | -22 | 65 | N/A | Z= 3.56 |
|  | 0 | R postcentral gyrus | 38 | -33 | 62 | N/A | Z= 3.49 |
|  | 0 | R subgyral | 23 | -46 | 57 | N/A | Z= 3.34 |
|  | 0 | L superior parietal lobule | -25 | -46 | 58 | N/A | Z= 4.34 |
|  | 0 | L postcentral gyrus | -30 | -24 | 44 | N/A | Z= 4.00 |
|  | 0 | L postcentral gyrus | -31 | -37 | 58 | N/A | Z= 3.37 |
|  | 0 | L medial frontal gyrus | -13 | 36 | 35 | N/A | Z= 4.07 |
|  | 0 | L medial frontal gyrus | -7 | 30 | 36 | N/A | Z= 3.93 |
|  | 0 | L superior frontal gyrus | -13 | 46 | 35 | N/A | Z= 3.37 |
|  | 0 | L inferior temporal gyrus | -55 | -18 | -18 | N/A | Z= 3.89 |
|  | 0 | L anterior cingulate | -5 | 36 | 18 | N/A | Z= 3.77 |
|  | 0 | L paracentral lobule | -2 | -37 | 59 | N/A | Z= 3.68 |
|  | 0 | L superior temporal gyrus | -41 | -31 | 15 | N/A | Z= 3.63 |
|  | 0 | L transverse temporal gyrus | -35 | -35 | 11 | N/A | Z= 3.46 |
|  | 0 | L insula | -38 | -24 | 18 | N/A | Z= 3.10 |
|  | 0 | L Middle temporal gyrus | -42 | -56 | 24 | N/A | Z= 3.54 |
|  | 0 | L insula | -35 | 18 | 4 | N/A | Z= 3.41 |
|  | 0 | inferior occipital gyrus | -29 | -93 | -5 | N/A | Z= 3.38 |
|  | 0 | L cuneus | -22 | -93 | -1 | N/A | Z= 3.29 |
|  | 0 | L cingulate gyrus | -5 | 7 | 36 | N/A | Z= 3.26 |
| *Li et al, 2019* | 0 | L Inferior frontal gyrus | -28.5 | 19.5 | -22.5 | 172 voxels | T= -5.18 |
|  | 0 | L Rectus | -16.5 | 15 | -15 | 99 voxels | T= -4.53 |
|  | 0 | R Rectus | 19.5 | 19.5 | -13.5 | 181 voxels | T= -5.22 |
|  | 0 | R Middle temporal gyrus | 49.5 | -21 | -19.5 | 127 voxels | T= -4.96 |
|  | 0 | L Middle temporal gyrus | -55.5 | -19.5 | -16.5 | 99 voxels | T= -4.91 |
|  | 0 | L Inferior frontal gyrus | -48 | 42 | -7.5 | 27 voxels | T= -3.92 |
|  | 0 | L Middle temporal gyrus | -48 | -37.5 | 7.5 | 65 voxels | T= -4.90 |
|  | 0 | R Superior temporal gyrus | 49 | -33 | 15 | 44 voxels | T= -4.13 |
|  | 0 | R superior frontal gyrus | 13.5 | 46.5 | 30 | 14 voxels | T= -4.40 |
|  | 0 | R Precuneus | 13.5 | -55.5 | 42 | 102 voxels | T= -4.79 |
|  | 0 | R Postcentral gyrus | 45 | -21 | 48 | 92 voxels | T= -4.68 |
|  | 1 | R caudate | 18 | -7.5 | 19.5 | 243 voxels | T= -4.37 |
| *Raznahan et al, 2012* | 1 | L Superior Frontal Gyrus | -13 | 65 | 21 | N/A | T= 5.1 |
|  | 1 | L Superior temporal sulcus | -48 | -38 | 2 | N/A | T= 3.8 |
|  | 1 | R Superior Frontal Gyrus | 9 | 62 | 27 | N/A | T= 3.9 |
|  | 1 | R Middle Frontal Gyrus | 39 | 55 | -2 | N/A | T= 3.3 |
|  | 1 | R Superior Temporal Sulcus | 59 | -26 | 1 | N/A | T= 3.5 |
|  | 1 | R Rostral Intraparietal Sulcus | 32 | -62 | 53 | N/A | T= 4 |
| *Osipowicz et al, 2015* | 0 | L/R Thalamus | 3 | -42 | -6 | 280,690k | T= 5.7 |
|  | 0 | L Inferior Frontal Gyrus | -35 | 57 | -15 | 3032k | T= 4.7 |
|  | 0 | R Orbitofrontal Gyrus | 8 | 63 | -21 | 2641k | T= 4.5 |
|  | 0 | R Amygdala | 18 | 2 | -27 | 1971k | T= 4.2 |
|  | 0 | L Parahippocampal gyrus | -20 | 0 | -32 | 2002k | T= 4.1 |
| *Waiter et al, 2004* | 1 | L superior frontal gyrus | -23 | 60 | 11 | N/A | Z= 4.54 |
|  | 1 | R fusiform Gyrus | 41 | -59 | -5 | N/A | Z= 4.20 |
|  | 1 | R medial frontal gyrus | 2 | 10 | -16 | N/A | Z= 4.11 |
|  | 1 | L middle temporal gyrus | -44 | 5 | -26 | N/A | Z= 3.53 |
|  | 1 | R posterior cingulate | 14 | -50 | 18 | N/A | Z= 3.40 |
|  | 1 | L superior frontal gyrus | -15 | 6 | 67 | N/A | Z= 3.38 |
|  | 1 | L superior temporal gyrus | -32 | 8 | -20 | N/A | Z= 3.33 |
|  | 1 | R superior temporal gyrus | 43 | -54 | 30 | N/A | Z= 3.25 |
|  | 1 | L lingual gyrus | -22 | -89 | -1 | N/A | Z= 3.23 |
|  | 1 | L inferior frontal gyrus | -47 | 19 | -6 | N/A | Z= 3.22 |
|  | 1 | L middle frontal gyrus | -31 | 48 | -5 | N/A | Z= 3.78 |
|  | 1 | L inferior occipital gyrus | -37 | -71 | -1 | N/A | Z= 3.15 |
|  | 1 | L parahippocampal gyrus | -21 | -16 | -16 | N/A | Z= 3.09 |
|  | 0 | R thalamus | 17 | -15 | 17 | N/A | Z= 3.18 |
| *Kosaka et al, 2010* | 0 | R Insula (anterior + posterior) | 44 | -12 | 10 | 90k | T= 4.91 |
|  | 0 | R Insula (anterior) | 44 | 4 | 2 | 15k | T= 4.45 |
|  | 0 | R Insula (anterior) | 46 | 14 | -4 | 12k | T= 4.38 |
|  | 0 | R Inferior Frontal Gyrus | 58 | 20 | 10 | 26k | T= 5.25 |
|  | 0 | R Inferior Parietal | 30 | -38 | 58 | 2k | T= 4.20 |
| *Richter et al, 2015* | 0 | L Superior frontal gyrus | -7.3 | 60.3 | 18.5 | 152.36 mm^3 | p= 0.0057 |
|  | 0 | L superior frontal gyrus | -10.6 | -4.5 | 46.6 | 72.25 mm^3 | ^ |
|  | 0 | L superior frontal gyrus | -9.3 | 31.2 | 38.5 | 164.33 mm^3 | ^ |
|  | 0 | L rostral part of midddle frontal gyrus | -22.7 | 49.8 | 19.3 | 138.67 mm^3 | p= 0.0006 |
|  | 0 | L rostral part of middle frontal gyrus | -45.6 | 24.7 | 33.8 | 44.22 mm^3 | ^ |
|  | 0 | L caudal part of middle frontal gyrus | -36.6 | 10.3 | 34.7 | 104.59 mm^3 | p= 0.0043 |
|  | 0 | L middle part of orbitofrontal gyrus | -0.4 | 41.9 | -20.4 | 70.84 mm^3 | N/A |
|  | 0 | L middle part of orbitofrontal gyrus | -9.4 | 53.2 | -1.9 | 44.67 mm^3 | N/A |
|  | 0 | L orbital part of inferior frontal gyrus | -40.8 | 35.6 | -12.9 | 210.98 mm^3 | N/A |
|  | 0 | L Opercular part of inferior frontal gyrus | -49.4 | 12 | 12.3 | 131.75 mm^3 | N/A |
|  | 0 | L Superior temporal gyrus | -52.4 | -34 | 7.8 | 36.43 mm^3 | p= 0.0339 |
|  | 0 | L Insula | -34 | 12.2 | -2.4 | 133.73 mm^3 | N/A |
|  | 0 | L Parahippocampal gyrus | -18.7 | -33.4 | -12.6 | 6.76 mm^3 | N/A |
|  | 0 | L parahippocampal gyrus | -21.2 | -20.4 | -26 | 7.22 mm^3 | N/A |
|  | 0 | L Fusiform Gyrus | -38.3 | -48.3 | -20.1 | 23.54 mm^3 | N/A |
|  | 0 | L Inferior Temporal Gyrus | -46.7 | -52.6 | -11.4 | 86.54 mm^3 | p= 0.0021 |
|  | 0 | L Precuneus | -4.9 | -64.2 | 28.4 | 286.24 mm^3 | N/A |
|  | 0 | L Precuneus | -10.2 | -44.1 | 41.5 | 27.62 mm^3 | N/A |
|  | 0 | R Precuneus | 5.9 | -40.2 | 41.4 | 22.80 mm^3 | N/A |
|  | 0 | L Inferior Parietal Gyrus | -31.5 | -74.3 | 19.2 | 87.13 mm^3 | p= 0.0134 |
|  | 0 | L Lateral Occipital Gyrus | -26.7 | -91.4 | 13.3 | 19.43 mm^3 | N/A |
|  | 0 | L Lateral Occipital Gyrus | -43.5 | -78.3 | 2.8 | 26.81 mm^3 | N/A |
|  | 0 | L Lingual Gyrus | -21 | -67.8 | 0.7 | 12.24 mm^3 | N/A |
|  | 0 | L Cuneus | -10.3 | -95 | 10 | 357.35 mm^3 | N/A |
|  | 1 | R Caudal part of Middle Frontal Gyrus | 41.2 | 19.7 | 43.9 | 7.96 mm^3 | p= 0.0125 |
| *D'Mello et al, 2015* | 0 | R Crus I/II | 24 | -70 | -39 | 170k | T= 3.91 |
|  | 0 | R Lingual Gyrus | 11 | -84 | -15 | 157k | T= 4.23 |
|  | 0 | R Angular Gyrus | 40 | -78 | 40 | 79k | T= 3.53 |
|  | 1 | L Posterior Cingulate/Precuneus | -8 | -46 | 18 | 80k | T= 4.11 |
|  | 1 | R Superior Frontal Gyrus | 20 | 41 | 39 | 82k | T= 3.92 |
|  | 1 | L Middle Occipital Gyrus | -39 | -82 | 12 | 137k | T= 3.74 |
| *David et al, 2013* | 0 | L Frontopolar Gyrus | -20 | 66 | 0 | 1425k | T= 6.84 |
|  | 0 | R Parahippocampal Gyrus | 36 | -43 | -5 | 542k | T= 6.19 |
|  | 0 | R Inferior Frontal Gyrus | 33 | 34 | -3 | 579k | T= 5.30 |
|  | 0 | L Hippocampus | -36 | -16 | -15 | 2256k | T= 5.17 |
|  | 0 | L Superior Temporal Sulcus | -68 | -40 | 6 | 651k | T= 5.15 |
|  | 0 | R Angular Gyrus | 40 | -75 | 45 | 430k | T= 4.63 |
|  | 0 | L Insula | -44 | 6 | -2 | 958k | T= 4.46 |
| *Zielinksi et al, 2012* | 1 | L SMA | -14 | -15 | 62 | N/A | T= 3.55 |
|  | 1 | L SMA | -10 | -6 | 58 | N/A | T= 2.07 |
|  | 1 | L Superior Frontal | -20 | -6 | 58 | N/A | T= 1.95 |
|  | 1 | R Inferior Frontal | 36 | 27 | -10 | N/A | T= 1.92 |
|  | 1 | R SMA | 12 | -3 | 55 | N/A | T= 1.86 |
|  | 1 | L Precentral Gyrus | -49 | 1 | 40 | N/A | T= 1.81 |
|  | 1 | R Caudate | 2 | 16 | -2 | N/A | T= 1.73 |
|  | 0 | R Mid Frontal | 42 | 39 | -14 | N/A | T= 4.86 |
|  | 0 | L Mid Frontal | -28 | 55 | 7 | N/A | T= 4.2 |
|  | 0 | R Sup Frontal | 27 | 55 | 12 | N/A | T= 4.03 |
|  | 0 | L Medial Sup Frontal | -7 | 51 | 14 | N/A | T= 3.61 |
|  | 0 | L Medial Sup Frontal | -11 | 56 | 5 | N/A | T= 2.87 |
|  | 0 | L Medial Sup Frontal | -5 | 30 | 40 | N/A | T= 2.36 |
|  | 0 | R Mid Temporal Pole | 45 | 9 | -2 | N/A | T= 3.49 |
|  | 0 | L Mid Frontal | -37 | 46 | 17 | N/A | T=3.19 |
|  | 0 | L Mid Frontal | -31 | 44 | 26 | N/A | T= 3.05 |
|  | 0 | L Sup Frontal | -18 | 47 | 34 | N/A | T= 2.24 |
|  | 0 | L Sup Frontal | -24 | 41 | 35 | N/A | T= 2.09 |
|  | 0 | R Sup Frontal | 16 | 54 | 18 | N/A | T= 2.16 |
|  | 0 | L Inf Frontal | -49 | 32 | 18 | N/A | T= 1.81 |
|  | 0 | L Sup Temporal Pole | -43 | 1 | -26 | N/A | T= 1.74 |
|  | 0 | L Mid Temporal Pole | -40 | 12 | -29 | N/A | T= 1.69 |
|  | 0 | L Inf Frontal | -44 | 27 | -15 | N/A | T= 1.68 |
|  | 0 | R Inf Frontal | 51 | 23 | -2 | N/A | T= 1.68 |
|  | 0 | L Inf Frontal | -43 | 24 | -15 | N/A | T= 1.66 |
|  | 1 | R Precuneus | 14 | -73 | 40 | N/A | T= 3.61 |
|  | 1 | R Precuneus | 2 | -70 | 30 | N/A | T= 3.58 |
|  | 1 | L Mid Occipital | -30 | -78 | 35 | N/A | T= 3.51 |
|  | 1 | R Mid Occipital | 46 | -74 | 28 | N/A | T= 3.48 |
|  | 1 | R Angular Gyrus | 52 | -66 | 25 | N/A | T= 2.86 |
|  | 1 | R Sup Temporal | 57 | -24 | -3 | N/A | T= 2.15 |
|  | 1 | R Inf Temporal | 59 | -54 | -11 | N/A | T= 2 |
|  | 1 | R Mid Temporal Pole | 57 | -58 | -3 | N/A | T= 1.9 |
|  | 1 | L Mid Occipital | -48 | -76 | 16 | N/A | T= 1.98 |
|  | 1 | R Sup Temporal | 48 | -13 | 1 | N/A | T= 1.71 |
|  | 1 | R Mid Temporal Pole | 53 | -61 | 15 | N/A | T= 1.67 |
|  | 0 | L Inf Parietal | -56 | -46 | 39 | N/A | T= 3.79 |
|  | 0 | L Inf Parietal | -49 | -47 | 46 | N/A | T= 3.59 |
|  | 0 | L Postcentral gyrus | -46 | -34 | 4 | N/A | T= 3.51 |
|  | 0 | R Precentral | 61 | 1 | 24 | N/A | T= 3.5 |
|  | 0 | R Inf Parietal | 49 | -36 | 53 | N/A | T= 2.5 |
|  | 0 | R Postcentral gyrus | 50 | -27 | 53 | N/A | T= 2.16 |
|  | 0 | L Precuneus | -6 | -58 | 57 | N/A | T= 2.42 |
|  | 0 | L Sup Parietal | -19 | -56 | 54 | N/A | T= 2.19 |
|  | 0 | L Precuneus | -1 | -50 | 53 | N/A | T= 1.93 |
|  | 0 | L Inf Parietal | -51 | -30 | 48 | N/A | T= 2.3 |
|  | 0 | L Mid Temporal Pole | -57 | -56 | -6 | N/A | T= 2.12 |
| *Pereira et al, 2018* | 0 | R Posterior Cerebellum | 33 | -55 | -53 | 1804 voxels | T= 4.24 |
|  | 0 | L Fusiform Gyrus | -44 | -54 | -8 | 259 voxels | T= 4.58 |
|  | 0 | R Anterior Cerebellum | 14 | -60 | -30 | 347 voxels | T= 4.45 |
|  | 0 | L Cingulate Gyrus | -6 | -13 | 7 | 1562 voxels | T= 4.43 |
|  | 0 | R Cingulate Gyrus | 8 | -9 | 42 | ^ | T= 4.25 |
|  | 0 | L Paracentral Lobule | -8 | -9 | 45 | ^ | T= 4.25 |
|  | 0 | R Middle Frontal Gyrus | 32 | 53 | -14 | 263 voxels | T= 4.42 |
|  | 0 | L Claustrum | -38 | -10 | 3 | 642 voxels | T= 4.12 |
|  | 0 | R Medial Frontal Gyrus | 12 | 51 | 1 | 170 voxels | T= 4.06 |
|  | 0 | L Parahippocampal gyrus | -15 | -18 | -26 | 66 voxels | T= 3.89 |
|  | 0 | L Lentiform nucleus | -18 | -9 | -9 | 121 voxels | T= 3.85 |
|  | 0 | L Amygdala | -26 | -7 | -14 | ^ | T= 3.74 |
|  | 0 | L Postcentral gyrus | -6 | -42 | 70 | 73 voxels | T= 3.72 |
|  | 0 | L Posterior Cerebellum | -30 | -58 | -48 | 77 voxels | T= 3.69 |
|  | 0 | R Superior Frontal Gyrus | 1 | 60 | 30 | 37 voxels | T= 3.57 |
|  | 0 | R Cingulate gyrus | 18 | 33 | 22 | 32 voxels | T= 3.49 |
|  | 1 | R Posterior Cerebellum | 45 | -45 | -38 | 96 voxels | T= 3.93 |
|  | 1 | Brainstem | -2 | -37 | -27 | 42 voxels | T= 3.52 |
| *Scheel et al, 2011* | 0 | L Middle Temporal Gyrus | -49.7 | -60.9 | 9.3 | 1435.5 mm^3 | p= 0.0400 |
| *Itahashi et al, 2015* | N/A - No VBM findings | N/A | N/A | N/A | N/A | N/A | N/A |
| **AN MRI** | **GM Increase** | **Region** | **MNI/TAL Coordinates** | | | **Cluster Extent** | **Z/T/p value** |
|  | *0=decrease, 1=increase* |  | *x* | *y* | *z* | **(mm^3)/k** |  |
| *Nickel et al. 2018* | 0 | L superior frontal gyrus | -16 | -10 | 65 | 33k | Z= 4.18 |
|  | 0 | L superior frontal gyrus | -22 | 24 | 37 | 43k | Z= 4.07 |
|  | 0 | R superior frontal gyrus | 17 | 17 | 60 | 205k | Z- 4.66 |
|  | 0 | R superior frontal gyrus | 21 | -6 | 60 | 43k | Z= 4.17 |
|  | 0 | L Middle frontal gyrus | -38 | 5 | 39 | 56k | Z= 4.19 |
|  | 0 | R middle frontal gyrus | 30 | 8 | 53 | 87k | Z= 4.21 |
|  | 0 | L Posterior medial frontal gyrus | -6 | 17 | 64 | 34k | Z= 4.13 |
|  | 0 | L inferior frontal gyrus (pars triangularis) | -54 | 23 | 17 | 32k | Z= 4.12 |
|  | 0 | L middle temporal gyrus | -40 | -62 | 20 | 174k | Z= 4.35 |
|  | 0 | L superior parietal gyrus | -15 | -40 | 43 | 8k | Z= 4.06 |
|  | 0 | L midcingulate cortex | -12 | -52 | 38 | 179k | Z= 4.48 |
|  | 0 | R midcingulate cortex | 13 | -30 | 46 | 160k | Z= 4.67 |
|  | 0 | R precuneus | 17 | -70 | 46 | 376k | Z= 4.90 |
|  | 0 | R precuneus | 8 | -55 | 44 | 288k | Z= 4.69 |
|  | 0 | R insular lobe | 38 | 23 | 9 | 39k | Z= 4.17 |
| *Björnsdotter et al. 2018* | 0 | R putamen | 33 | 3 | 2 | 240 voxels | T= 3.21 |
|  | 0 | L/R Hypothalamus | -3 | 3 | -9 | 82 voxels | T= 3.38 |
| *Martin Monzon et al. 2017* | 0 | L amygdala | -18.5 | 1.5 | -24 | 70.926 voxels | T= 10.49 |
|  | 0 | L caudate nucleus | -13.5 | 9 | 19.5 | ^ | ^ |
|  | 0 | L ACC | -3 | 36 | 0 | ^ | ^ |
|  | 0 | L MCC | -10.39 | -15.85 | 38.24 | ^ | ^ |
|  | 0 | L/R PCC | 0 | -46.83 | 17.57 | ^ | ^ |
|  | 0 | L insular cortex | -39 | -1.5 | 9 | ^ | ^ |
|  | 0 | R insular cortex | 45 | 1.5 | 7.5 | ^ | ^ |
|  | 0 | L hippocampus | -30 | -40.5 | 1.5 | ^ | ^ |
|  | 0 | L occipital cortex | -9 | -94.5 | -9 | ^ | ^ |
|  | 0 | R occipital cortex | 19.5 | -99 | -9 | ^ | ^ |
|  | 0 | L OFC | -36 | 36 | -12 | ^ | ^ |
|  | 0 | R OFC | 36 | 39 | 10.5 | ^ | ^ |
|  | 0 | L parietal association cortex | -26.19 | -76.87 | 31.09 | ^ | ^ |
|  | 0 | R parietal association cortex | 29.53 | -76.87 | 31.09 | ^ | ^ |
|  | 0 | L precuneus | -4.5 | -57 | 51 | ^ | ^ |
|  | 0 | R Precuneus | 7.5 | -51 | 64.5 | ^ | ^ |
|  | 0 | L DLPFC | 45.63 | 34.71 | 22.62 | ^ | ^ |
|  | 0 | R DLPFC | 40.82 | 48.56 | 21.08 | ^ | ^ |
|  | 0 | LmPFC | 15 | 57 | -12 | ^ | ^ |
|  | 0 | R MPFC | 16.5 | 58.5 | -10.5 | ^ | ^ |
|  | 0 | L thalamus | -4.5 | -11.54 | 14.98 | ^ | ^ |
|  | 0 | R Thalamus | 7.5 | -3 | 6 | ^ | ^ |
|  | 0 | R amygdala | 25.5 | 1.5 | -21 | 604 voxels | T= 4.21 |
|  | 0 | R caudate nucleus | 15 | 9 | 21 | 523 voxels | T= 5.60 |
|  | 0 | R hippocampus | 30 | -37.5 | 6 | 525 voxels | T= 4.75 |
| *Kohmura et al. 2017* | 0 | R superior temporal gyrus | 48 | -36 | 12 | 1156 voxels | T= 4.51 |
|  | 0 | R middle temporal gyrus | 51 | -36 | -11 | ^ | T= 4.39 |
|  | 0 | R superior temporal gyrus | 54 | -40 | 9 | ^ | T= 4.37 |
|  | 0 | R middle cingulate gyrus | 3 | 14 | 37 | 15.109 voxels | T= 5.79 |
|  | 0 | R anterior cingulate | 0 | -16 | 45 | ^ | T= 5.49 |
|  | 0 | R anterior cingulate | 0 | 53 | 10 | ^ | T= 5.49 |
|  | 0 | L angular grus | -39 | -64 | 43 | 1530 voxels | T= 5.44 |
|  | 0 | L superior Occipital gyrus | -29 | -79 | 39 | ^ | T= 3.97 |
|  | 0 | L angular gyrus | -45 | -70 | 33 | ^ | T= 3.85 |
|  | 0 | L middle temporal gyrus | -57 | -19 | -9 | 1144 voxels | T= 5.10 |
|  | 0 | L middle temporal gyrus | -59 | -12 | -24 | ^ | T= 4.53 |
|  | 0 | L middle temporal gyrus | -56 | 3 | -23 | ^ | T= 4.38 |
|  | 0 | L middle frontal gyrus | -26 | 44 | 19 | 877 voxels | T= 5.08 |
|  | 0 | L central perculum | -56 | -3 | 3 | 1815 voxels | T= 4.88 |
|  | 0 | L inferior frontal gyrus (pars triangularis) | -54 | 6 | 18 | ^ | T= 4.72 |
|  | 0 | L parietal lobule | -29 | -31 | 48 | ^ | T= 4.39 |
| *Fuglset et al. 2016* | 0 | L superior parietal gyrus | -25.1 | 58.6 | 58.1 | 1766.95 mm^2 | p< 0.05 |
|  | 0 | R superior parietal gyrus | 18.8 | -71.1 | 44.1 | 2402.59 mm^2 | p< 0.05 |
|  | 0 | R inferior parietal gyrus | 41.1 | -71.6 | 35.5 | 2218.18 mm^2 | p< 0.05 |
|  | 0 | R superior frontal gyrus | 23.1 | 0.5 | 62.2 | 1674.74 mm^2 | p< 0.05 |
| *Seitz et al. 2015* | 0 | R inferior parietal lobe | 47 | -59.6 | 44.2 | 1163.43 mm2 | T= -7.33 |
|  | 0 | R insula | 35.2 | -14.8 | 19.3 | 4000.62 mm2 | T= -6.43 |
|  | 0 | R posterior cingulate | 5.2 | -33.7 | 40.9 | 5098.36 mm2 | T= -6.22 |
|  | 0 | R superior temporal gyrus | 53.2 | -28 | 0.6 | 1018.33 mm2 | T= -5.22 |
|  | 0 | R caudal middle frontal | 39.2 | 9 | 54.7 | 578.52 mm2 | T= -4.94 |
|  | 0 | R lingual | 21.1 | -47.9 | -5.2 | 3241.71 mm2 | T= -4.89 |
|  | 0 | R middle temporal gyrus | 54.1 | -59.2 | 4.9 | 2073.21 mm2 | T= -4.88 |
|  | 0 | R rostral middle frontal | 31.2 | 47.2 | 7.4 | 186.56 mm2 | T= -4.43 |
|  | 0 | R post central | 44.1 | -23.8 | 60.2 | 1475.07 mm2 | T= -4.28 |
|  | 0 | R supramarginal | 55.1 | -36.2 | 17.6 | 744.85 mm2 | T= -3.90 |
|  | 0 | R lateralorbitofrontal | 32.4 | 33.3 | -8.4 | 454.6 mm2 | T= -3.90 |
|  | 0 | R superiortemporal | 48.9 | 8.2 | -23.7 | 159.82 mm2 | T= -3.78 |
|  | 0 | Rsuperior frontal | 10.9 | 23.4 | 56.1 | 292.11 mm2 | T= -3.76 |
|  | 0 | R lateraloccipital | 25.2 | -97.5 | 0 | 448.36 mm2 | T= -3.51 |
|  | 0 | R rostralmiddle frontal | 23.3 | 43.9 | 23.4 | 164.71 mm2 | T= -3.44 |
|  | 0 | R superiorparietal | 19.2 | -81.5 | 38.2 | 1036.91 mm2 | T= -3.32 |
|  | 0 | R middle temporal gyrus | 64.1 | -34.7 | -13.3 | 306.12 mm2 | T= -3.28 |
|  | 0 | R supramarginal | 53.2 | -21.4 | 17.8 | 157.61 mm2 | T= -3.13 |
|  | 0 | R inferior parietal | 44 | -65.9 | 25.9 | 111.2 mm2 | T= -3.09 |
|  | 0 | R caudal middle frontal | 33.6 | 7.4 | 26.4 | 187.42 mm2 | T= -3.05 |
|  | 0 | R precentral | 55 | 2.5 | 22.8 | 518.5 mm2 | T= -3.04 |
|  | 0 | R medial orbitofrontal | 11.4 | 32.7 | -18.5 | 197.49 mm2 | T= -2.89 |
|  | 0 | R lateral orbitofrontal | 17.6 | 38.3 | -18.2 | 86.42 mm2 | T= -2.69 |
|  | 0 | R pericalcarine | 17 | -92.8 | 1.6 | 213.28 mm2 | T= -2.58 |
|  | 0 | R supramarginal | 56 | -29.3 | 43.7 | 33.52 mm2 | T= -2.30 |
|  | 0 | R inferior parietal | 47.3 | -71.4 | 9.2 | 121.89 mm2 | T= -2.18 |
|  | 0 | R supramarginal | 51.9 | -45.1 | 31.4 | 26.92 mm2 | T= -2.18 |
|  | 0 | R superior parietal gyrus | 23 | -60.3 | 39.1 | 34.14 mm2 | T= -2.16 |
|  | 0 | R precentral | 38.5 | -13.1 | 56.9 | 44.59 mm2 | T= -2.13 |
|  | 0 | R precentral | 22.4 | -6.9 | 49.4 | 12.54 mm2 | T= -2.065 |
|  | 0 | R parahippocampal | 23.2 | -25.9 | -17.9 | 6.9 mm2 | T= -2.064 |
|  | 0 | R inferior parietal | 34.6 | -67.4 | 40 | 16.82 mm2 | T= -1.97 |
|  | 0 | R lateral occipital | 17.4 | -95.2 | 14.2 | 14.78 mm2 | T= -1.92 |
|  | 0 | L paracentral | -9.8 | -34.8 | 53.4 | 13475.25 mm2 | T= -7.56 |
|  | 0 | L superior parietal gyrus | -13.3 | -76.5 | 45.6 | 4814.63 mm2 | T= -6.58 |
|  | 0 | L middle temporal gyrus | -59.5 | -54.7 | 3.1 | 3980.24 mm2 | T= -6.44 |
|  | 0 | L postcentral | -50.4 | -20.5 | 55.1 | 1209.99 mm2 | T= -5.57 |
|  | 0 | L fusiform | -32.8 | -33.5 | -24 | 263.1 mm2 | T= -3.56 |
|  | 0 | L insula | -36.2 | -16.2 | 20.3 | 154.85 mm2 | T= -3.39 |
|  | 0 | L superior temporal | -51.4 | 6.3 | -14 | 434.32 mm2 | T= -3.066 |
|  | 0 | L supramarginal | -55.9 | -41.9 | 42 | 398.32 mm2 | T= -3.03 |
|  | 0 | L lateral occipital | -28.4 | -93.3 | -2.7 | 213.25 mm2 | T= -2.88 |
|  | 0 | L precentral | -49.6 | -7.7 | 36.7 | 148.06 mm2 | T= -2.74 |
|  | 0 | L superior temporal | -56.1 | -12.5 | 1.7 | 235.93 mm2 | T= -2.71 |
|  | 0 | L fusiform | -30 | -71.4 | -8.6 | 329.18 mm2 | T= -2.60 |
|  | 0 | L lateral orbitofrontal | -15.4 | 44.7 | -18.3 | 223.55 mm2 | T= -2.58 |
|  | 0 | L caudal middle frontal | -39.2 | 5.5 | 49.8 | 80.33 mm2 | T= -2.53 |
|  | 0 | L banks sts | -52.1 | -36 | 1.4 | 64.46 mm2 | T= -2.33 |
|  | 0 | L isthmus cingulate | -4.4 | -41.9 | 28.3 | 85.36 mm2 | T= -2.33 |
|  | 0 | L inferior parietal lobe | -33.6 | -65.2 | 41.3 | 41.66 mm2 | T= -2.27 |
|  | 0 | L insula | -36.4 | -17.4 | 9.5 | 79.92 mm2 | T= -2.25 |
|  | 1 | L precentral | -31.9 | -24.6 | 51.7 | 38.61 mm2 | T= 2.22 |
|  | 0 | L superior parietal gyrus | -36 | -51.9 | 59.8 | 44.29 mm2 | T= -2.21 |
|  | 0 | L superior parietal gyrus | -14.2 | -87.8 | 24.5 | 86.3 mm2 | T= -2.19 |
|  | 0 | L medial orbitofrontal | -7.2 | 29.8 | -17.1 | 158.61 mm2 | T= -2.14 |
|  | 0 | L fusiform | -41.1 | -52.2 | -18.7 | 59.22 mm2 | T= -2.093 |
|  | 0 | L inferior parietal lobe | -43.5 | -57.1 | 38.6 | 29 mm2 | T= -2.077 |
|  | 0 | L supramarginal | -59.7 | -49.5 | 27.6 | 37.05 mm2 | T= -2.057 |
|  | 0 | L superior temporal | -48.2 | -30.7 | 3 | 19.37 mm2 | T= -1.91 |
|  | 0 | L lateral occipital | -23.6 | -86 | 12.9 | 13.62 mm2 | T= -1.90 |
|  | 0 | L inferior parietal lobe | -38.8 | -63 | 29 | 10.01 mm2 | T= -1.89 |
|  | 0 | L fusiform | -36.2 | -14.5 | -29.5 | 17.18 mm2 | T= -1.88 |
|  | 0 | L middle temporal gyrus | -43.2 | 7 | -35 | 12.6 mm2 | T= -1.87 |
|  | 0 | L lateral occipital | -13.7 | -101.7 | 3.9 | 16.89 mm2 | T= -1.87 |
|  | 0 | L superior parietal gyrus | -30.4 | -56.8 | 36.9 | 1.46 mm2 | T= -1.80 |
| *Fujisawa et al. 2015* | 0 | L inferior frontal gyrus (pars triangularis) | -52 | 30 | 24 | 434 voxels | Z= 4.40 |
|  | 0 | R inferior frontal gyrus | 58 | 30 | 20 | 225 voxels | Z= 4.04 |
| *Van Opstal et al. 2015* | 0 | L cingulate cortex | -4 | -16 | 48 | 237 voxels | p< 0.05 |
|  | 0 | R cingulate cortex | 8 | 22 | 30 | 231 voxels | p< 0.05 |
| *D'Agata et al. 2015* | 0 | L cerebellum (VI) | -24 | -54 | -26 | 9459 mm3 | p= 0.0032 |
|  | 0 | R cerebellum (VI) | 24 | -50 | -26 | 7049 mm3 | p= 0.0036 |
|  | 0 | L Supplementary Motor Area | -4 | -16 | 64 | 4772 mm3 | p= 0.0028 |
| *Bär et al. 2015* | 0 | R posterior cingulate cortex | 8 | -43 | 42 | 3708 mm2 | p= 0.011 |
|  | 0 | R precuneus | 5 | -61 | 43 | ^ | p= 0.018 |
|  | 0 | L midcingulate cortex | -5 | -26 | 44 | ^ | p= 0.026 |
|  | 0 | R midcingulate cortex | 12 | -26 | 41 | ^ | p= 0.023 |
|  | 0 | R supplementary motor area | 2 | 0 | 56 | ^ | p= 0.020 |
| *Frank et al. 2013* | 1 | L middle orbitofrontal cortex | -33 | 60 | -9 | 189 | T= 4.21 |
|  | 1 | R fusiform gyrus | 20 | -34 | -15 | 137 | T= 5.43 |
|  | 1 | R fusiform gyrus | 27 | -10 | -36 | 107 | T= 4.88 |
|  | 1 | L fusiform gyrus | -32 | -7 | -41 | 100 | T= 4.26 |
|  | 1 | R hippocampus | 29 | -27 | -8 | 113 | T= 4.74 |
|  | 1 | L hippocampus | -33 | -19 | -11 | 74 | T= 4.01 |
|  | 1 | R insula | 41 | 5 | 7 | 69 | T= 4.06 |
|  | 1 | R parahippocampal gyrus | 18 | -10 | -29 | 579 | T= 6.41 |
|  | 1 | L parahippocampal gyrus | -21 | -13 | -27 | 116 | T= 4.42 |
|  | 1 | L gyrus rectus | -5 | 30 | -26 | 19 | T= 3.99 |
|  | 1 | L gyrus rectus | -9 | 30 | -26 | 25 | T= 3.95 |
|  | 1 | L gyrus rectus | -6 | 53 | -24 | 41 | T= 4.19 |
|  | 1 | R Hippocampus | 29 | -21 | -20 | 208 | T= 5.60 |
|  | 1 | R Parahippocampal Gyrus | 29 | -25 | -20 | 237 | T= 5.74 |
|  | 1 | R Middle temporal gyrus | 50 | -31 | -6 | 82 | T= 5.98 |
|  | 1 | L superior temporal gyrus | -48 | -33 | 22 | 98 | T= 3.86 |
| *Lázaro et al. 2013* | N/A - No VBM findings | | N/A | N/A | N/A | N/A | N/A |
| *Amianto et al. 2013* | 1 | R paracentral/precuneus | 12 | -38 | 52 | 659k | p< 0.001 |
|  | 0 | L SMA | -6 | -17 | 64 | 1229k | p< 0.001 |
|  | 0 | L Cerebellum Crus I | -36 | -48 | -36 | 657k | p= 0.001 |
|  | 0 | R Cerebellum Crus I | 52 | -70 | -28 | 441k | p= 0.001 |
|  | 0 | R thalamus | 10 | -18 | 0 | 356k | p< 0.001 |
|  | 0 | R Precentral | 22 | -18 | 48 | 115k | p= 0.001 |
|  | 0 | R Occipital inferior | 36 | -70 | -4 | 113k | p< 0.001 |
|  | 0 | L Fusiform | -24 | -22 | -32 | 104k | p< 0.001 |
|  | 0 | R Supramarginal | 39 | -30 | 38 | 73k | p= 0.001 |
|  | 0 | L Hippocampus | -28 | -6 | -22 | 67k | p= 0.001 |
| *Brooks et al. 2011* | 0 | L cerebellum | -3 | -52 | -16 | 2.61 cm3 | p< 0.002 |
|  | 0 | L/R Parahippicampal gyrus | 24 | -36 | 4 | 0.77 cm3 | p< 0.002 |
|  | 0 | R Anterior Insula | 31 | 13 | -12 | 0.52 cm3 | p< 0.002 |
|  | 0 | L Fusiform Gyrus | -25 | -93 | 4 | 0.52 cm3 | p< 0.002 |
|  | 0 | R Posterior Cingulate | 4 | -45 | 32 | 0.42 cm3 | p< 0.002 |
|  | 1 | R DLPFC | 34 | 45 | 28 | 0.59 cm3 | p< 0.002 |
| *Friederich et al. 2012* | 0 | R insula/amygala | 37 | 3 | -16 | 1188 voxels | T= 5.29 |
|  | 0 | R amygdala/putamen | 28 | 1 | -12 | ^ | T= 5.18 |
|  | 0 | L Hippocampus | -8 | 8 | -13 | 1784 voxels | T= 6.19 |
|  | 0 | L Amygdala/Putamen | -18 | 0 | -12 | ^ | T= 5.34 |
|  | 0 | L Temporal pole sup./Insula | -38 | -1 | -18 | ^ | T= 4.37 |
|  | 0 | R Anterior cingulate | 6 | -15 | 41 | 7098 voxels | T= 6.14 |
|  | 0 | R supplementary motor area | -6 | 4 | 54 | ^ | T= 5.44 |
|  | 0 | R anterior cingulate | 11 | -25 | 45 | ^ | T= 5.26 |
| *Boghi et al. 2011* | 0 | R opercular roland | 43 | -15 | 22 | 17348 voxels | T= 6.14 |
|  | 0 | R parietal inferior lobe | 47 | -54 | 44 | ^ | T= 5.50 |
|  | 0 | R fusiform gyrus | 40 | -62 | -14 | 6409 voxels | T= 6.11 |
|  | 0 | R head caudate | 5 | 10 | 6 | 1707 voxels | T= 5.58 |
|  | 0 | R hypothalamus | 2 | -3 | -11 | 100 voxels | T= 5.46 |
|  | 0 | L fusiform | -28 | -53 | -8 | 2315 voxels | T=5.28 |
|  | 0 | L parahippocampal gyrus | -32 | -42 | -3 | ^ | T= 4.87 |
|  | 0 | R Cerebellum (VIII) | 34 | -43 | -43 | 5707 voxels | T= 4.92 |
|  | 0 | L Caudate nucleus | -3 | 5 | 6 | 1341 voxels | T= 4.92 |
|  | 0 | R Paracentral lobule | 0 | -37 | 66 | 1296 voxels | T= 4.39 |
|  | 0 | R Supplementary motor area | 2 | -30 | 53 | ^ | T= 4.18 |
|  | 0 | R precentral | -42 | 0 | 33 | 120 voxels | T= 4.30 |
|  | 0 | R mid occipital | 44 | -86 | 7 | 314 voxels | T= 4.12 |
|  | 0 | L mid temporal | -47 | -52 | 7 | 255 voxel | T= 4.12 |
|  | 0 | L hypothalamus | -3 | -2 | -10 | 48 voxels | T= 4.11 |
|  | 0 | R frontal inferior | 40 | 26 | 11 | 103 voxels | T= 4.00 |
|  | 0 | L Cerebellum (VI) | -16 | -59 | -26 | 199 voxels | T= 3.84 |
|  | 0 | Precentral R | 55 | -11 | 46 | 67 voxels | T= 3.77 |
|  | 0 | L Superior Parietal | -19 | -68 | 41 | 57 voxels | T= 3.74 |
| *Joos et al. 2010* | 0 | R rostral anterior cingulum | 12 | 54 | -1 | 699 voxels | T= 4.29 |
|  | 0 | L Dorsal anterior cingulum | -1 | 19 | 32 | 641 voxels | T= 3.93 |
|  | 0 | R frontal operculum | 55 | -2 | 13 | 4737 voxels | T= 4.91 |
|  | 0 | L precuneus | -1 | -59 | 31 | 1072 voxels | T= 4.05 |
|  | 0 | L temporoparietal cortex | -37 | -65 | 31 | 3503 voxels | T= 4.53 |
|  | 0 | L parietal cortex | -50 | -34 | 60 | 600 voxels | T= 3.86 |
|  | 0 | R cerebellum | 42 | -73 | -56 | 38 voxels | T= 3.76 |
| *Castro-Fornieles et al. 2009* | 0 | R Superior Temporal | 68 | -32 | 3 | 13320 voxels | T= 8.36 |
|  | 0 | L Precuneus | 32 | -50 | 69 | 35593 voxels | T= 8.14 |
|  | 0 | L Inferior Parietal | -45 | -48 | 61 | 3078 voxels | T= 6.45 |
| *Leppanen et al. 2019* | 0 | L SPC | -22.9 | -64.2 | 29.7 | 168.61 mm2 | Z= -5.01 |
|  | 0 | L Paracentral | -14.4 | -41 | 67.1 | 133.06 mm2 | Z= -4.21 |
|  | 1 | L ACC | -5.6 | 34.4 | 2.6 | 130.60 mm2 | Z= 4.46 |
|  | 0 | R SPC | 11.7 | -52.2 | 64.9 | 189.52 mm2 | Z= -4.78 |
|  | 0 | R LOC | 21.3 | -88.4 | 18.2 | 189.29 mm2 | Z= -5.98 |
|  | 0 | R precuneus | 18.1 | -70.8 | 35.7 | 158.01 mm2 | Z= -5.92 |
| *Fonville et al. 2014* | 0 | R Cerebellum | 26 | -56 | -34 | 3229 voxels | T= 4.61 |
|  | 0 | R temporal occipital fusiform cortex | 28 | -50 | -16 | ^ | ^ |
|  | 0 | L cerebellum | -28 | -56 | -36 | 1573 voxels | T= 5.02 |
|  | 0 | L lateral occipital cortex | -20 | -66 | 34 | 1144 voxels | T= 4.26 |
|  | 0 | L precuneus | -2 | -54 | 54 | ^ | ^ |
|  | 0 | R lateral occipital cortex | 30 | -90 | 28 | 733 voxels | T= 4.2 |
|  | 0 | R cuneus | 8 | -84 | 26 | ^ | ^ |
|  | 0 | L Lateral occipital cortex | -24 | -92 | 20 | 238 voxels | T= 4.109 |
|  | 0 | R Superior frontal gyrus | 20 | 12 | 46 | 225 voxels | T= 4.917 |
|  | 0 | R Precuneus | 12 | -68 | 24 | 183 voxels | T= 3.371 |
| *Suchan et al. 2010* | 0 | L Lateral Occipital Cortex | -48 | -66 | 9 | 45k | Z= 4.90 |
|  | 0 | L superior temporal gyrus | -56 | -50 | 15 | 19k | Z= 5.10 |
| *Gaudio et al. 2011* | 0 | Middle cingulate cortex | -7 | -41 | 31 | 17359 voxels | T= 7.11 |
|  | 0 | Precuneus | 0 | 54 | 40 | 7492 voxels | T= 6.46 |
|  | 0 | L/R Inferior Superior Parietal lobule | -38 | -48 | 43 | 6571 voxels | T= 6.08 |
| *Olivo et al. 2018* | N/A - No VBM findings | N/A | N/A | N/A | N/A | N/A | N/A |

**Supplementary Table 1. All ASD and AN MNI/Talairach coordinates used across literature for analysis.**

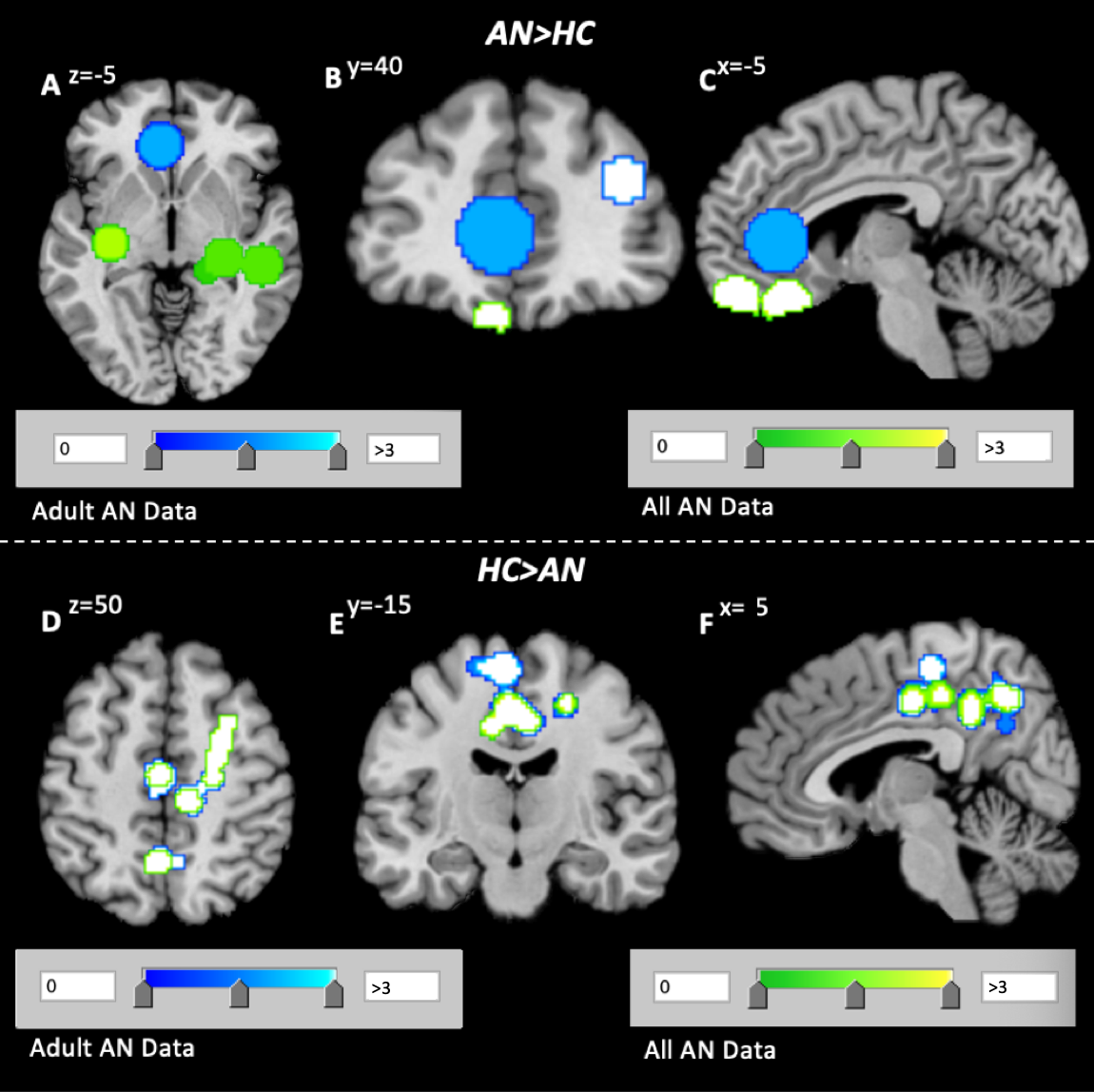
*Supplementary Figure 1. (ADULTS vs. ALL DATA)*

**Supplementary Figure 1.** Comparison of cluster results found in all AN data (green) compared to adult-only data (blue). Clusters are much more separate upon states of increased volume, while quite homogenous for clusters of decreased volume.

*Supplementary Table 2 ([ADULTS] AN>HC; AN<HC)*

| **[ADULTS]**  **AN>HC** | | **Region** | **BA** | **MNI Coordinates** | | | **Volume (mm^3^)** | **ALE score** | **P** | **Z** |
| --- | --- | --- | --- | --- | --- | --- | --- | --- | --- | --- |
|  |  |  |  | *x* | *y* | *z* |  |  |  |  |
| ***C1*** | ***R Paracentral Lobule*** | | ***5*** | **12** | **-38** | **52** | **13,944** | **0.009** | **2.24E-05** | **4.08** |
| ***C2*** | ***L Anterior Cingulate*** | | ***24*** | **-6** | **34** | **2** | **12,176** | **0.01** | **4.69E-06** | **4.43** |
| ***C3*** | ***R Orbitofrontal Cortex (MFG)*** | | ***10*** | **38** | **52** | **20** | **8,704** | **0.01** | **2.68E-05** | **4.04** |
| **AN<HC** | |  |  |  | | |  |  |  |  |
| ***C1*** | ***R Cingulate Gyrus*** | | ***31*** | **12** | **-26** | **44** | **17,768** | **0.02** | **5.06E-07** | **4.89** |
| *C1* | *L Orbitofrontal Cortex (MFG)* | | *6* | -4 | -16 | 64 |  | 0.017 | 6.02E-06 | 4.37 |
| *C1* | *L Paracentral Lobule* | | *31* | -2 | -16 | 46 |  | 0.015 | 2.74E-05 | 4.03 |
| *C1* | *R Precuneus* | | *7* | 6 | -58 | 44 |  | 0.013 | 1.12E-04 | 3.69 |
| *C1* | *R Cingulate Gyrus* | | *31* | 8 | -44 | 40 |  | 0.012 | 1.89E-04 | 3.55 |
| *C1* | *L Precentral Gyrus* | | *6* | -16 | -10 | 64 |  | 0.01 | 5.92E-04 | 3.24 |
| *C1* | *L Precuneus* | | *7* | -2 | -54 | 54 |  | 0.01 | 6.45E-04 | 3.21 |
| *C1* | *L Cingulate Gyrus* | | *31* | -4 | -26 | 44 |  | 9.9E-03 | 7.78E-04 | 3.16 |
| *C1* | *R Paracentral Lobule* | | *6* | 4 | -24 | 58 |  | 9.4E-03 | 1.0E-03 | 3.03 |
| *C1* | *R Cingulate Gyrus* | | *31* | 22 | -18 | 48 |  | 9.3E-03 | 1.3E-03 | 3.00 |
| *C1* | *L Cingulate Gyrus* | | *31* | 0 | -60 | 32 |  | 8.3E-03 | 3.4E-03 | 2.71 |

**Supplementary Table 1.** Aberrant GMV in the AN adult subgroup relative to age-matched controls, including adult studies only (nAN=15). Data is presented in order of significance, with coordinate sets in bold comprising of cluster centres.

[***Abbreviations:*** *L – left; R – right; MFG – Medial frontal gyrus; BA – Brodmann Area; C - Cluster*]
